# Unobtrusive inference of diurnal rhythms from smartphone data

**DOI:** 10.1101/2025.04.14.25325807

**Authors:** Loran Knol, Mindy K. Ross, Anisha Nagpal, Andrew P. Burns, Zachery D. Morrissey, Faraz Hussain, Tory A. Eisenlohr-Moul, Christian F. Beckmann, Alex Leow, Andre F. Marquand

## Abstract

Diurnal rhythms are an integral feature of psychopathology but difficult to measure at scale. Smartphones are ubiquitous and therefore uniquely positioned to measure such rhythms non-invasively and continuously. Here, we propose a digital phenotyping framework to quantify diurnal rhythms. We use it to predict sleep duration from smartphone typing dynamics and analyse rhythm phase during time zone transitions with a clinical outpatient sample and a year-long longitudinal data set.

## Introduction

Diurnal rhythms are a key feature of human biology and behaviour, and are disrupted in many forms of psychopathology^1^, including major depressive disorder^2^, schizophrenia^3^, and bipolar disorder^4^. For example, a person’s preferred waking and sleeping time (chronotype), is associated with depressive and manic symptoms such that an evening chronotype (preferring to go to bed late) confers added risk^5,6^. Such rhythms are also apparent in cognition in that different facets of cognition fluctuate uniquely over time, for example during the day, in a highly individualised manner. They depend on an individual’s chronotype and interact with factors such as stress, motivation, and the quality of the previous night’s sleep^7–9^. Moreover, long-term disruption of diurnal rhythm, for instance due to shift work or frequent exposure to jet lag, impacts both physical health and cognitive performance^10,11^.

However, studying these rhythms outside the lab and over long timescales remains challenging with traditional methods. For sleep, polysomnography is considered the gold standard but also the most burdensome^12^. Self-report is less burdensome but subjective and prone to recall bias and attrition^13–15^. Similarly, objectively measuring fluctuations in cognition requires repeated administration of tests, which is logistically challenging and burdensome. Recent advancements in smartphone and wearable technology, however, have allowed researchers to passively measure or approximate different aspects of human behaviour and physiology via digital phenotyping^16^. Wearables such as the Apple Watch, Fitbit, and Oura ring have been used to predict biological sleep features using, e.g., actigraphy^17–25^, heart rate variability^24–27^, and skin temperature^17,22,26^. Smartphones, however, offer a unique advantage for indexing behaviour, as they are ubiquitous in modern life and most people interact with their phone throughout the day. This means that studies employing smartphone-based measurements need less additional hardware, making such studies considerably cheaper than those that need to acquire digital wearables.

Over the last eight years, we have developed and validated a leading smartphone-based platform, BiAffect, for unobtrusive monitoring of behavioural and mental health through keyboard typing dynamics and continuous sensor data.^28–39^ BiAffect features an iOS app that replaces the default iPhone keyboard and collects typing speed, autocorrect rate, backspace rate, and phone movement and orientation while typing. These typing dynamics are associated with executive processing speed and show a diurnal pattern in optimal typing performance^32,33^. We have also shown that this approach is sensitive to behavioural fluctuations linked to clinically relevant mental states, predicting symptoms such as depressed mood, mania, and anhedonia^28–30,32,38,39^. Meanwhile, research on other smartphone platforms has shown that measurements such as GPS, screen state, accelerometery, and even typing also predict sleep characteristics^40–44^. This warrants the development of a framework that integrates these measures, offering a comprehensive perspective on behavioural and cognitive fluctuations that complements the physiological insights provided by digital wearables.

Here, we provide a framework to detect and quantify diurnal rhythms unobtrusively based on smartphone behaviour – leveraging the smartphone’s capability to passively index cognition over extended timescales – with the potential for extensions to other data. Core considerations while designing this framework were the management of missing data, which is a key challenge in digital phenotyping^45,46^, and scalability to potentially large numbers of diverse data types from various devices. Our method provides a fully automated approach to detect diurnal rhythms by creating a low-rank approximation of a collection of digital phenotyping modalities using graph-regularised singular value decomposition^47^, preserving the data’s inherent temporal structure. Our approach is predicated on the assumption that most people have relatively stable 24-hour rhythms, allowing periods of missing data to be imputed by referencing adjacent points in time and corresponding times on other days. This method results in a quantitative, time-resolved measure of device activity that can be used to infer diurnal rhythms. An advantage of this approach is that no assumptions are required about when people are most active during the day, allowing for unrestricted chronotypes and non-contiguous sleep episodes.

As a first step, we use this approach to map the rest-activity cycle non-invasively and unobtrusively over extended periods of time, predicting sleep duration from smartphone typing behaviour in a clinical sample. Next, we demonstrate the utility of our method for understanding the desynchronisation between diurnal behaviour and the external clock by analysing the diurnal phase of a year’s worth of BiAffect data of author AL, during which she frequently travelled internationally, as well as the phase of the aforementioned clinical sample. Our method provides a starting point for the analysis of diurnal variations in behavioural patterns collected from dense digital phenotyping data.

### Diurnal framework

Our framework for the analysis of diurnal patterns in digital phenotyping data is based on graph-regularised singular value decomposition (GRSVD; see Supplementary Note 3 for details). A prerequisite for this approach is that each data modality is aggregated into regular bins – in our case, hours – and arranged into day-by-bin matrices (Figure 1A). We extracted hourly aggregates of four BiAffect modalities: 1) Median alphanumeric inter-key delay, which is an inverse measure of typing speed, 2) the total number of key presses, 3) upright typing rate, which measures the proportion of typing sessions in which the phone was oriented upwards, and 4) movement rate, which measures the proportion of typing sessions in which the phone was non-stationary.

**Figure 1:**
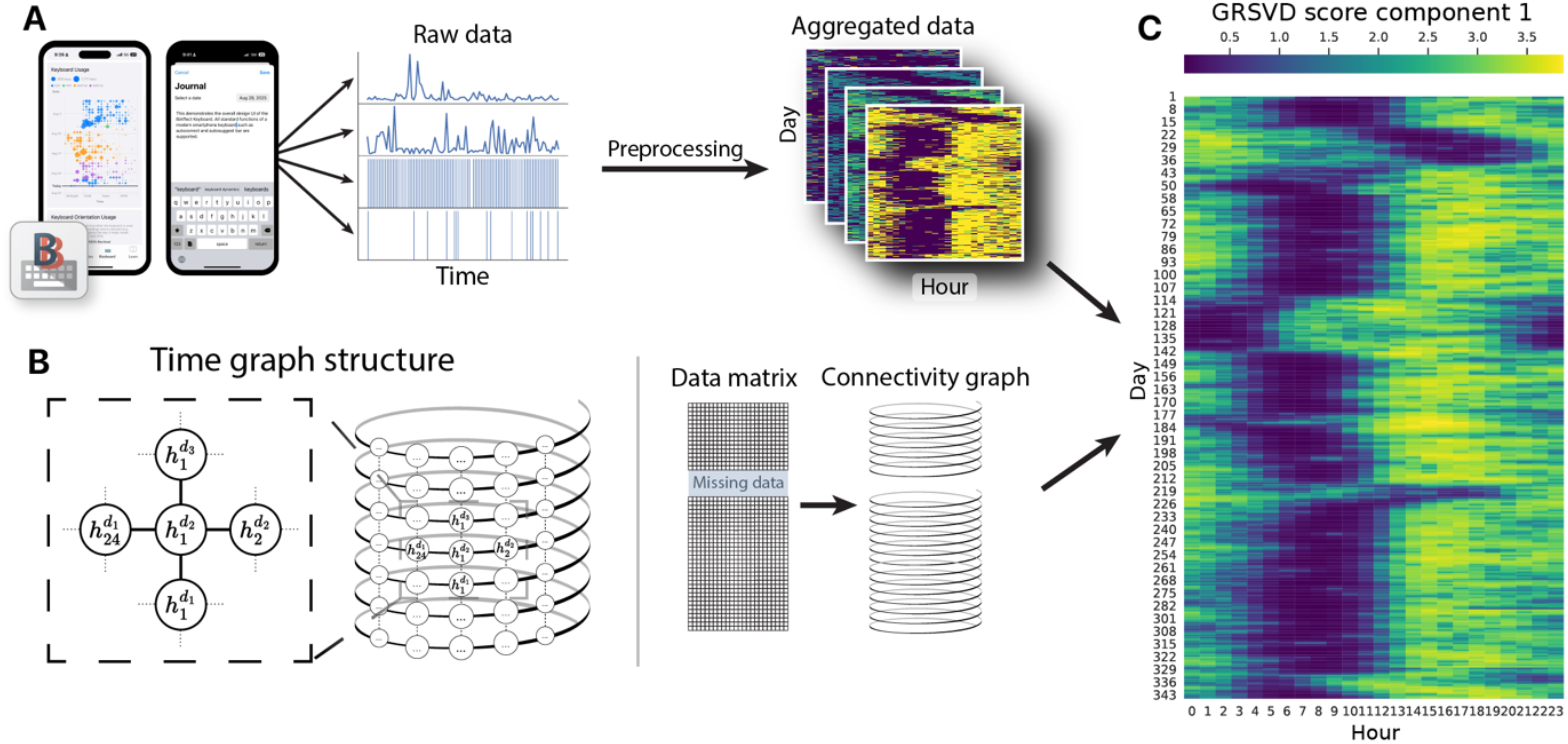
Overview of the graph-regularised SVD. **A** Keyboard typing dynamics are collected by the BiAffect app, preprocessed, and aggregated into hourly bins. For every typing modality, the aggregates are arranged into day-by-hour matrices. See Supplementary Figure 1 for a closer look at the BiAffect keyboard and dashboard. **B** Graph of the time structure underlying the hourly bins. Every hour 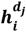 is connected to the previous hour 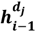 and subsequent hour 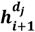. This also means that the final hour of a given day is connected to the first hour of the next day. Similarly, every hour is also connected to the same hour on the previous day 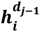 and the one on the subsequent day 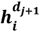. Conceptually, this leads to a graph that resembles a spiral with crosslinks. Missing days of data result in a disjoint graph, preventing information ffow to non-adjacent nodes. **C** The data matrices and time graph are fed into the graph-regularised SVD. This figure shows the first component of the decomposition, where the matrix values (GRSVD scores) can be interpreted as a general measure of typing activity.

Next, we constructed a graph that specified which matrix cells are neighbours due to the underlying temporal structure (Figure 1B). More specifically, we connected every hour to the preceding and subsequent hours and to its corresponding hours on the preceding and subsequent day. These connections encoded our assumption that a person has a sleep-wake rhythm that slowly fluctuates during the day and is approximately constant across days, enabling the GRSVD to selectively impute missing data by borrowing information from adjacent hours.

The GRSVD takes both the data matrices and the time graph to produce a dimensionality reduction in which the resulting components have been smoothed based on the graph connectivity (Figure 1C). The amount of regularisation (smoothing) depends on a hyperparameter, which was optimised by hand (Supplementary Figure 8). The first component reflects the dominant mode of variation across all input modalities and, for our data, revealed distinct sections of activity and inactivity that captured the user’s diurnal activity patterns. We used this component in all downstream analyses. Other components are shown in Supplementary Figure 13.

Further processing was required to extract relevant features. To predict sleep duration from smartphone data, we determined an appropriate threshold for binarisation of GRSVD typing activity scores into ‘sleeping’ or ‘wakeful’ and then counted the number of hours spent sleeping (Figure 2A). We additionally quantified daily typing rhythm phase by calculating a weighted circular mean of the observations of each day (row) of the GRSVD matrix (Figure 3A). See Supplementary Note 5 for details. All analysis source code is freely available on GitHub: https://github.com/Valkje/graph-svd.

**Figure 2:**
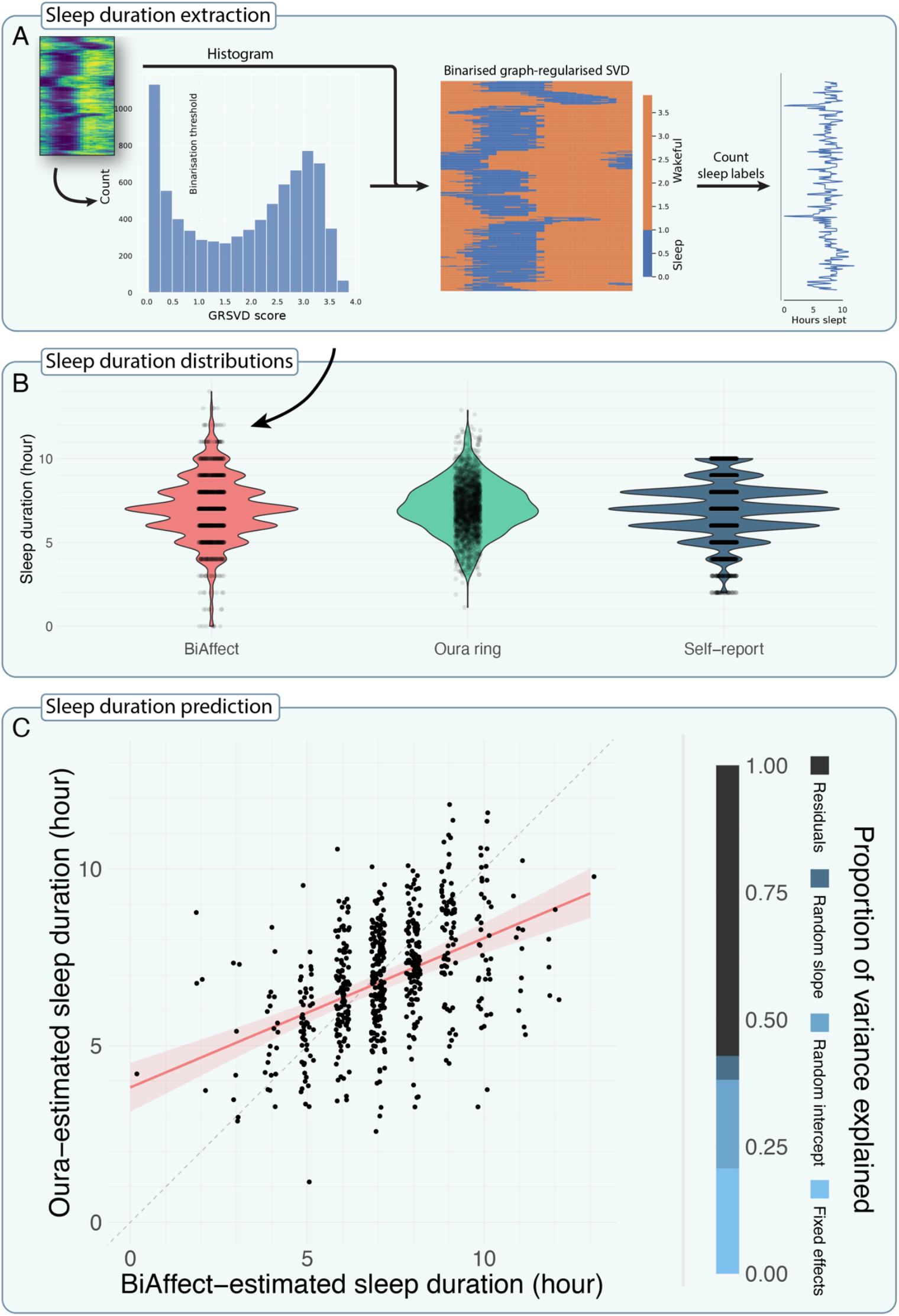
Validation of the graph-regularised SVD method on the CLEAR-3 dataset. **A** Algorithm overview for extracting sleep duration from a GRSVD matrix. First, a histogram is made of the matrix values from Figure 1C. The dashed line gives the manually determined binarisation threshold. Matrix cells with a value below the binarisation threshold are classified as sleep, everything else is considered wakeful. Sleep labels are counted for every day, resulting in a daily sleep duration prediction. **B** Violin plots of estimated sleep duration for BiAffect (total n = 1471), the smart ring (n = 227S), and self-report (n=7S70). Jittered data points are overlaid to show the data type (continuous or discrete). Typing-derived sleep estimates showed larger right-skew than the other distributions. These extreme data points corresponded to days for which small number of data were collected, possibly due to underutilisation of the phone. In addition, the ring data only concern sleep duration during bedtime, while BiAffect also measures daytime naps if they are present. BiAffect and the ring show a significant Spearman’s rank correlation r_s_ = 0.4C (n = C04, p < 0.0001). **C** Mixed model predictions of ring-derived sleep duration. Data are plotted as jittered points on top of the regression line. The dashed line is the y=x diagonal. The shaded band represents the S5% CI. The vertical bar graph indicates which proportion of the total variance is explained by which part of the model. The fixed effects, random intercepts, and random slopes respectively explained 20.7%, 17.4%, and 4.7% of the total variance, leaving 57.2% unexplained. The model displayed a root mean square error (RMSE) of 1.17 and a mean absolute error (MAE) of 0.S0; only incorporating the fixed effects yielded an RMSE of 1.47 and an MAE of 1.1C. A baseline model with only an intercept and participant-level random intercepts showed an RMSE of 1.32 and an MAE of 1.00. Excluding random slopes resulted in a significant decrease in model fit according to a likelihood ratio test (χ^2^(2) = 12.78, p = 0.0017). Subsequently excluding the fixed effects further decreased model fit (χ^2^(1) = 111.7C, p < 0.0001).

**Figure 3:**
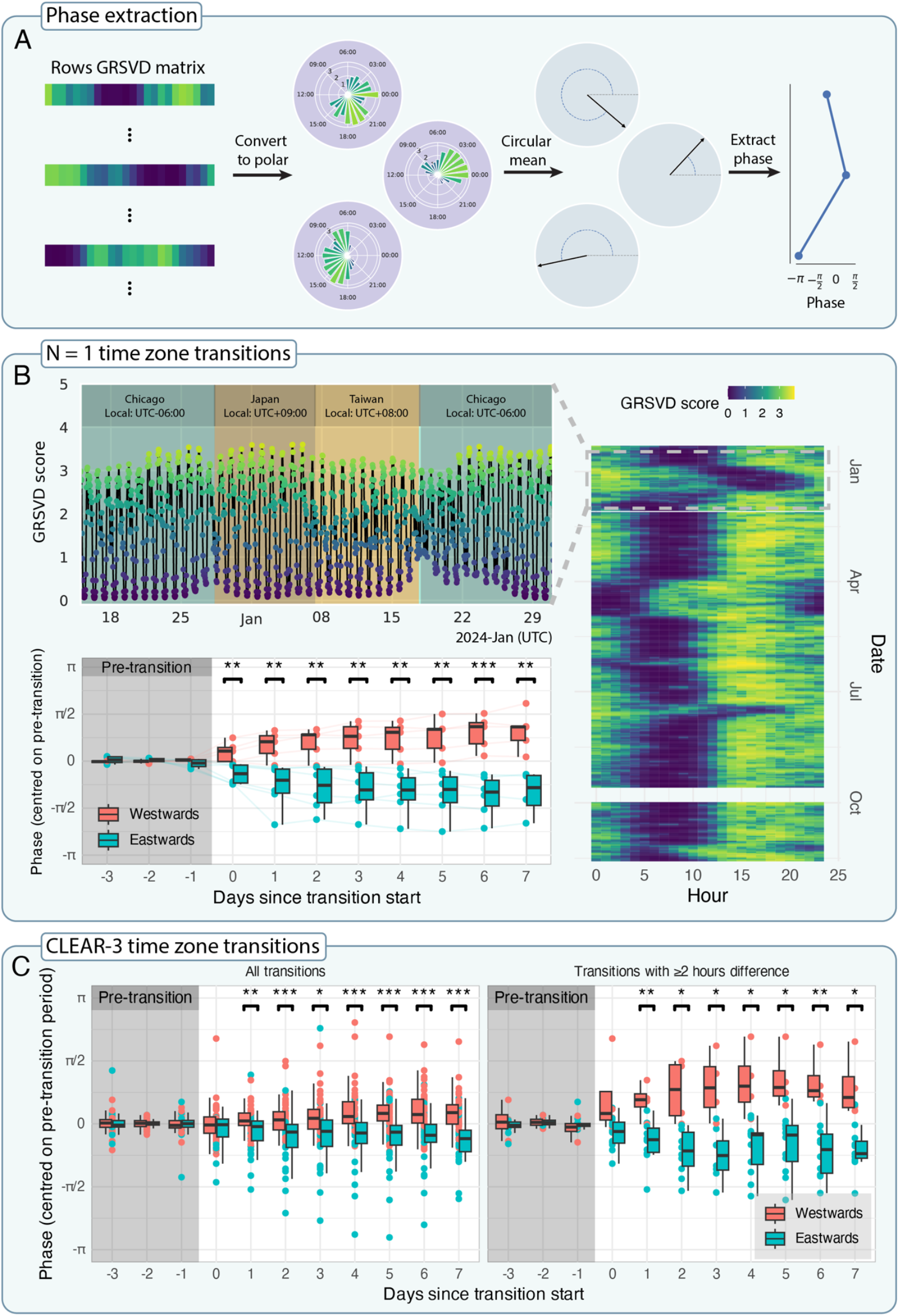
Phase analyses for two different samples. **A** Algorithm for extracting phase from the GRSVD matrix. Every matrix row is reinterpreted in polar coordinates, where the angle is given by the hour in UTC and the vector length corresponds to the GRSVD score. From these, we calculated weighted circular means, which indicate the time of peak activity, and extracted the angles of those means. These angles represent the phase of the daily typing activity rhythm. **B** N=1 analysis of nearly a year’s worth of typing data from author AL. Collection started on December 10, 2023, resulting in a nearly continuous data stream from that moment until we pulled the data on November 18, 2024, except for a brief interruption at the end of September 2024 due to technical issues. AL made seventeen transitions, visiting nine time zones (not shown). Top-left: Line plot of the GRSVD scores plotted over time (in UTC) rather than in matrix form. Dates range from December 15, 2023 to January 31, 2024, corresponding to AL’s trip from Chicago to Japan and Taiwan. Background colour indicates the time zone. Bottom-left: Quantitative analysis of the time-zone transitions that lasted at least seven days. Phases are displayed from three days preceding the transition to seven days following the transition, coloured by direction of travel. All transition phases are centred within-transition on the average phase of their pre-transition periods (shaded grey rectangle). Box plots visually summarise the statistics of each travel direction, for each day. The central line shows the median, while the lower and upper hinges represent the first and third quartile, respectively. Whiskers reach no further than 1.5 times the inter-quartile range. Brackets above pairs of box plots indicate a significant difference between travel directions according to Welch’s t-tests (p < 0.05: *; p < 0.01: **; p < 0.001: ***). **C** Quantitative analysis of CLEAR-3 participant phases with time zone transitions. Figure characteristics are the same as in B, bottom-left panel. Left: All time-zone transitions. Right: Only transitions with at least a 2-hour difference. Test statistics for all Welch’s t-tests are given in Supplementary Table 2.

### Validation analysis

To validate our framework, we applied it to data from the CLEAR-3 trial, a randomised controlled crossover trial (NCT04112368) that investigated a hormonal intervention to prevent perimenstrual exacerbation of suicidal ideation in individuals assigned female at birth. The study was approved by the UIC Institutional Review Board and all participants provided informed consent. Deidentified participant data will be made available on reasonable request to the principal investigator of the CLEAR-3 trial, T.A. Eisenlohr-Moul Sleep duration prediction used only the baseline data of the trial (i.e., prior to the intervention and placebo), which consisted of at least one menstrual cycle. Phase analyses used data from the entire trial and included healthy controls.

The participants were encouraged to use BiAffect throughout the study and self-reported on sleep duration using an hourly Likert scale, with a maximal response of 10+ hours of sleep. A subset, not including healthy controls, also wore Oura Rings (Oura Health Ltd, Oulu, Finland), a digital wearable (smart ring) that estimates sleep duration using accelerometery, photoplethysmography (measuring blood volume changes in the skin using light), and body temperature. These are regarded as good proxies of sleep quality and have been shown to yield 89% to 96% accuracy in detecting wakefulness and sleep as validated against polysomnography^17,27,48^. The version of the Oura Ring used in this study (Generation 2) did not record daytime naps.

Finally, we augmented our phase analysis with almost a year’s worth of data from author AL, which contained several periods of global travel. Details about filtering, preprocessing, and aggregation of all data are given in Supplementary Note 2.

Mixed-effects models were used to predict sleep duration as derived from the smart ring^49^. To account for repeated measures within participants, we included random intercepts and slopes for the BiAffect-derived sleep duration predictor. This predictor was standardised (M = 0, SD = 1) across all observations. See Supplementary Note 4 for more information.

For our diurnal rhythm phase analysis, we tested whether our phase metric would be sensitive to the direction of time zone change by comparing phases of east- and westwards travel using Welch’s t-tests. Transitions were centred within-transitions on the three days leading up to the transition (pre-transition period) and required to persist for at least seven days.

### Smartphone keyboard data predict sleep duration in a clinical sample

To test our computational framework, we calculated the graph-regularised SVDs (GRSVDs) of the BiAffect keyboard dynamics for every CLEAR-3 participant (Supplementary Figure 4), allowing us to derive an estimate of their sleep duration (Figure 2A) and compare it to the smart ring data. We generated typing-based estimates for 38 participants with these smart rings (numbers of participants before filtering and merging of data streams are given in Supplementary Figure 3). Demographics are shown in Supplementary Table 1. The distribution of the typing-derived estimates showed a larger tail than the ring and the (truncated) self-report distributions (Figure 2B), and is discrete rather than continuous because it was calculated from a binarised matrix (Figure 2A). Otherwise, the distributions matched in shape, showing their mode around seven hours of sleep. The linear mixed-effects models showed that our BiAffect-derived estimate of sleep duration significantly predicted ring-derived sleep duration (n = 604, β = 0.74, p < 0.0001; Figure 2C), although it showed a tendency for overestimation. Results for different regularisation levels and predictions using self-report data are given in Supplementary Figures 9-12 and Supplementary Table 5. Overall, these findings show that our framework mapped the rest-activity cycle by significantly predicting sleep duration from typing data.

### Smartphone keyboard data track diurnal phase

Next, we demonstrate that our approach can automatically detect phase shifts in diurnal behaviour with two separate data sets. First, we utilise nearly a year’s worth of typing data from author AL to show how typing dynamics change during time zone transitions (Figure 3B). We plot her GRSVD scores over time for one of her trips to Asia, exposing daily oscillations that become disturbed around large time zone transitions, with a visible refractory period. Below that, we show that the phase we extract is sensitive to the direction of travel (east- or westwards; Figure 3B). Each point represents her phase during a specific transition for a specific day. The difference between east- and westwards travel phases becomes significant from the day the transition occurs. Westwards travel is accompanied by a phase increase because, when viewed from a common reference such as UTC+00:00, people living further west become active later during the day. Conversely, eastwards travel is accompanied by a phase decrease.

We replicate this analysis with a subset of the CLEAR-3 sample. Figure 3C shows 83 time zone transitions from 33 participants. Whether including all transitions or just those with at least a two-hour time zone difference, the difference between east- and westward transition phases becomes significant on the first day after the transition rather than the day itself. The increased spacing between the box plots in the case of ≥2-hour transitions suggests that larger transitions are accompanied by larger absolute phase deflections.

### Considerations

We proposed a new method for the analysis of diurnal fluctuations in unobtrusively collected digital phenotyping data. Our method transforms a collection of data modalities to a representation that exposes their principal components through graph-regularised SVD, respecting inherent time dynamics of the data, thus allowing selective imputation^50^. We reported two downstream analyses that quantified different properties of the user’s sleep-wake cycle. First, we proposed a method for predicting sleep duration, which we validated with smart ring data in a clinical, psychiatric female outpatient sample, which used the BiAffect keyboard throughout the study. Our BiAffect-derived sleep duration estimates significantly predicted total sleep time measured by the smart ring, although it should be noted that a non-trivial portion of the variance remained unexplained. Second, we introduced a method that extracts the phase of the daily device activity pattern and allows comparisons over extended periods of time. We applied this method to a year’s worth of author AL’s typing data as well as the outpatient sample described above, showing how it can reveal diurnal patterns which are not immediately evident from raw smartphone data.

Despite significantly predicting wearable-derived sleep duration, our typing-based estimates tended to variously over- and underestimate the actual number of hours slept. This is due to two conflicting goals of our analysis: Smoothing out hours without data due to missingness (i.e., no phone usage while awake), and preserving those without data due to sleep. We resolve this conflict by assuming regular sleep-wake rhythms and encoding that assumption into the GRSVD graph. Unfortunately, this assumption is no longer valid in extreme cases, such as when a participant only sleeps a few hours scattered throughout the day, or when a participant types very little (due to little phone use or preferring tapping and scrolling). Such patterns can result in over- and undersmoothing in the GRSVD matrix and, subsequently, extreme sleep duration estimations. For those clinical populations where regular sleep-wake cycles are uncommon, the regularisation hyperparameter could be re-tuned to provide optimised results.

The results of this analysis indicate that the typing-derived sleep durations form an imperfect approximation of the high-quality ring-derived data, but this does not mean that the typing-derived estimates are without value. More specifically, when employing wearables in the study design is not an option due to, e.g., their high cost, our smartphone-based method provides a cheap, highly unobtrusive alternative for predicting sleep in naturalistic settings (as evidenced by the year’s worth of data from author AL). We further emphasise that our framework provides an alternative and complementary perspective on diurnal rhythms by considering behaviour and cognition, and should by no means be regarded as a replacement for wearables like a smart ring or pure actigraphy devices, which primarily focus on physiology and have already proven to be robust and accurate^17,27,48^. Future research on mental health and digital phenotyping will most likely benefit from taking both perspectives into account.

We chose to demonstrate our framework with typing dynamics because of the known relationship between time-of-day and typing speed^32^, with the additional advantage that the BiAffect app has minimal impact on battery life. Other smartphone modalities would also have been suitable, however, as shown by several studies using ambient sound and light, GPS, screen state, or accelerometery to predict sleep^40–43^. Most used small sample sizes and self-report for validation, but recent studies feature larger samples^51,52^. A wide variety of machine learning models has been used in these studies, including Bayesian networks^43^, decision trees^42,43^, linear mixed-effects regression^44,52^, Bayesian models^51^, and rule-based classifiers^40^. Only a handful of these studies focus on the diurnal rhythm more generally by, for instance, identifying participant chronotypes^40^. None consider the effects of international travel.

Our computational framework allows the analysis of the desynchronisation of someone’s diurnal rhythms from the external environment, which we demonstrated with nearly a year of iPhone data from author AL, leveraging her frequent time zone transitions, and the CLEAR-3 sample. For AL, we highlighted several cases in which the transitions resulted in more irregular rhythms and phase shifts of the sleep-wake cycle. Some of these suggested larger disturbances for east-than westwards travel, which is consistent with recent research.^53^ Our quantitative phase analyses corroborated the notion that phase is sensitive to the direction of travel (east- or westwards). In sum, we believe our approach provides a useful metric for tracking the regularity and phase of someone’s diurnal rhythm.

Such a metric is useful because diurnal rhythms are important for our health. Shift workers whose schedules misalign with their chronotype (e.g., working an early morning shift as an evening person) show impaired cognition^54^, attenuated melatonin fluctuations, more sleep disturbance, and larger social jet lag^55,56^. The impact of these changes go beyond poor sleep as melatonin has an anti-metastatic effect, potentially explaining the higher rates of breast cancer among night-shift workers^11^. Similar results have been found for airline cabin crew^57,58^. Although eliminating shift work may not be possible in present-day society, maintaining a real-time index of someone’s diurnal rhythm could be helpful in monitoring its long-term disruption and facilitate intervention when those disruptions become severe. We plan to add this feature to the BiAffect dashboard to allow users to monitor their own rhythms. Future research could use our method to study, e.g., how people adapt to new time zones in a manner similar to how Willoughby et al. studied sleep and international travel with Oura ring data^53^. One could additionally investigate how smartphone rhythms respond to psychopathological disturbances, such as the onset of mood episodes in bipolar disorder, potentially allowing early warnings when someone deviates from their baseline rhythm.

One potential improvement to our framework would be to differentiate between different kinds of missing data, such as technical issues versus a lack of phone usage. Another option would be to extend our framework to group-level decompositions, such as in the work of Aledavood et al., who applied non-negative matrix factorisation to smartphone data to identify participant chronotypes^59^. Finally, future studies could apply our method to a broader range of data types from a variety of platforms, with the additional possibility to retain more than just one dimension of the GRSVD, potentially uncovering richer characterisations of diurnal dynamics if the data contains them. One example would be social media usage data, which have previously been shown to display diurnal rhythms^60^, but the generic nature of the GRSVD would allow the simultaneous analysis of many other modalities.

We conclude that our GRSVD is a promising tool for uncovering diurnal patterns in digital phenotyping data and has several appealing features. Our method makes few assumptions about the participant’s chronotype or the contiguity of their sleep, which contrasts with some of the existing methods^40,44^. Other advantages include its capability to incorporate the continuous and cyclical nature of time through a graph, allowing the method to selectively impute missing data using surrounding time points. Most important, however, is the method’s low cost compared to existing options such as smart rings and other wearables. In sum, this approach provides future clinical studies with a promising new method to index diurnal rhythms cheaply, unobtrusively, and over long timescales, as well as a principled way for individuals to monitor their own diurnal dynamics.

## Data Availability

Deidentified participant data will be made available on reasonable request to the principal investigator of the CLEAR-3 trial, T.A. Eisenlohr-Moul.

https://github.com/Valkje/graph-svd

## Acknowledgements

This study was funded by the European Research Council (101001118). TAE-M and AN received funding from the National Institute of Mental Health (RF1MH120843 and F30MH138058, respectively). CFB gratefully acknowledges funding from the Wellcome Trust Collaborative Award in Science 215573/Z/19/Z and the Netherlands Organization for Scientific Research Vici Grant No. 17854 and NWO-CAS Grant No. 012-200-013.

## Author contributions

LK: Conceptualisation; data curation; software; formal analysis; methodology; visualisation; writing – original draft. MKR: Conceptualisation; methodology; software. AN: Data curation; writing – review & editing. APB: Conceptualisation; methodology; software; writing – review & editing. ZDM: Conceptualisation; methodology; software; writing – review & editing. FH: Software; writing – review & editing. TAE-M: Data curation; funding acquisition; investigation; writing – review & editing. CFB: Funding acquisition; supervision; writing – review & editing. AL: Conceptualisation; funding acquisition; investigation; methodology; supervision; writing – review & editing. AFM: Conceptualisation; funding acquisition; methodology; supervision; writing – review & editing.

## Competing interests

CFB is founding director and shareholder of SBG Neuro Ltd. AL is a cofounder of KeyWise AI, previously served on the Medical Board for Buoy Health, and currently serves as a digital psychiatry advisor for Otsuka, USA. All other authors declare no competing interests.

## Notes

### Author Declarations

IRB of University of Illinois at Chicago gave ethical approval for this work

### Summary of Updates

Added quantitative analysis of phase metric during time zone transitions. Moved parts of the Methods to the Supplement. Renamed section headers. Added additional sensitivity analyses.

