## Supplement for "Unobtrusive inference of diurnal rhythms from smartphone data"

### Supplementary Note 1 – BiAffect

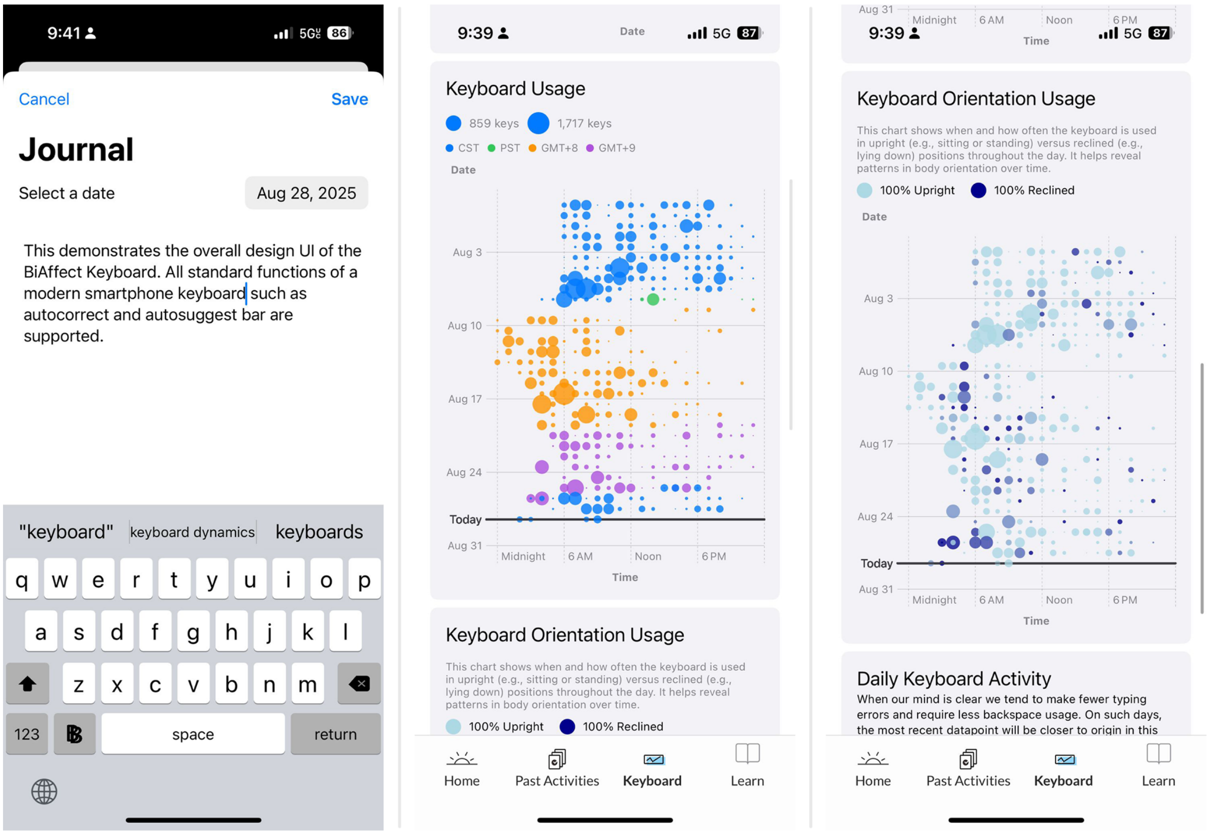

***Supplementary Figure 1: Screenshots from the BiAffect keyboard and dashboard.*** *Left: The BiAffect keyboard. Middle: A figure on the dashboard showing hourly aggregates of key press counts. Dot size represents number of key presses. Every row of dots corresponds to a day, and colours indicate time zone recorded by the phone. Right: Same as the middle panel, but now dots are coloured according to the proportion of time spent typing upright.*

Starting in 2018, the BiAffect project launched to develop a "fitness tracker for the brain". The project came about after author AL, being an avid amateur pianist, realised that having a good day positively impacted her piano play. She hypothesised that if her interaction with the piano keyboard would be indicative of her mood and cognition, this might also apply to the smartphone keyboard. Indeed, typing dynamics can be seen as the result of a psychomotoric process[1], and we have shown them to be correlated with mood, mania, anhedonia, and cognitive performance[2–8]. BiAffect's rationale is that the onset of psychopathology often comes with changes in cognition and smartphones are ideally positioned to sample that cognition because of their prevalence in our daily lives. BiAffect was originally geared towards bipolar disorder, where changes in mood and cognition are salient, but our studies have since expanded to include participants with other mental disorders as well as volunteers we call citizen scientists.

Through the phone's many sensors and on-device computing, BiAffect adopts a closed-loop experience design principle, attempting to be more than just a data-collecting research tool. More specifically, one of the goals of BiAffect is to empower the user by presenting their own data back to them. We do this through a dashboard that presents several visualisations of the user's typing behaviour (see Supplementary Figure 1). In doing this, we provide real-time feedback about each user's distinctive typing patterns to promote reflection and insight into their own psychomotoric state.

#### Supplementary Note 2 – Preprocessing

Preprocessing of BiAffect data largely followed the same procedure as in previous works[8,9], briefly summarised here. To limit the size of the typing data matrices, we aggregated data into hourly bins using several methods, yielding different features for analysis. First, we counted the total number of key presses for every hour. To constrain the range of key press counts and ensure positivity, we incremented them by one and took the natural log. We then calculated the inter-key delays (IKDs) between all alphanumeric key presses within a single typing session, defined as starting at the first key press and terminating when the participant either closed the keyboard or did not enter any key press for six seconds. Consequently, we took the median of these IKDs for every hour to derive an inverse measure of typing speed[9,2].
Accelerometer data were collected only when a typing session was active. Data were sampled at 10 Hz and filtered with a second-order bidirectional Butterworth filter with a cut-off of 4 Hz to remove high-frequency noise. We then classified typing sessions as moving or 'active' if more than 8% of the accelerometer samples had a magnitude lower than 0.95 or higher than 1.05 (a magnitude of 1 reflects the gravitational pull of the earth). To classify sessions as upright or lying down, we considered the median x- and z-values of the accelerometer samples: We would mark a session as upright if the median x-value was in-between -0.2 and 0.2 (inclusive) and the median z-value was below 0.1[9]. Otherwise, the session was considered as lying down. Finally, these Boolean session classifications were averaged per hour, giving rates of moving and upright sessions.
Since our method is contingent on the presence of enough data to distinguish between waking and sleeping hours, we applied additional processing steps before applying the graph-regularised SVD. We required participants to have at least, on average, 1) 20% of their daily hours filled with some data, 2) 50 key presses per hour, and 3) seven days of data, where a day was defined in UTC+00:00 rather than local time. We used UTC+00:00 because time zone transitions could cause overlap in data if converted to local time. Participants would be excluded from the data set if they did not meet these criteria. For remaining participants, we required at least 33% of every day to have at least either a) 1 key press for the phase analysis and b) 30 key presses for the mixed-effects models. We employed this distinction because we preferred to use as much data as possible for

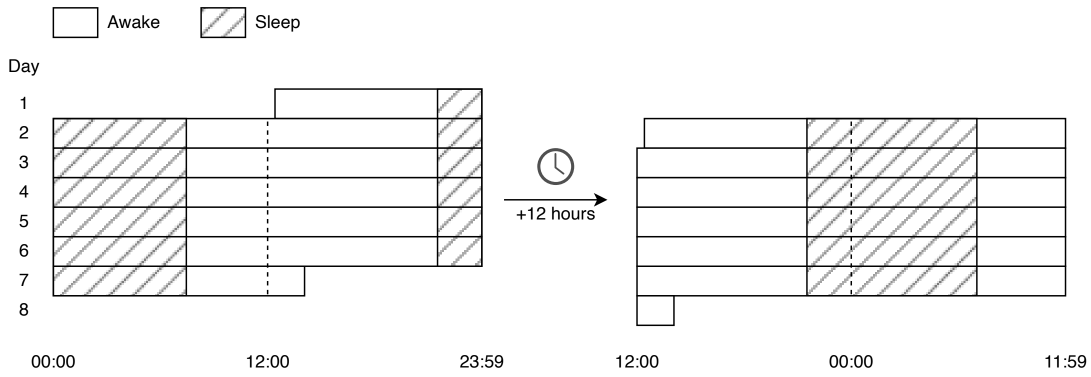

***Supplementary Figure 2: Schematic overview of the time shift applied during preprocessing.*** *Adding +12 hours to every data timestamp will ensure that every 24-hour period, represented as a row in our data matrix, has a complete night. Otherwise, every night could be dispersed over two rows. While this does not affect the outcome of the graph-regularised SVD, it does make it easier to count the number of hours slept in the time windows that the smart ring is most likely to use.*

phase analysis, whilst the predicted sleep durations markedly improved with more stringent filtering. Results for other thresholds are given in Supplementary Figure 6-Supplementary Figure 7 and Supplementary Table 3.
After running the GRSVD (described below), we excluded days with a time zone transition, converted all timestamps to local time, and right-shifted them with +12 hours to bring the nights to the centre of our 24-hour periods, which facilitates the calculation of the amount of time slept during those nights (see Supplementary Figure 2). From this point on, a 'day' is defined as the time period running from 12pm on one day to 11:59am on the next day.
All typing-derived features were scaled to unit variance (but not mean-centred) within participants, separately for the training and testing sets (described below).
This study used the Oura Ring Generation 2 to measure sleep duration. Oura ring total sleep time was derived according to proprietary algorithms. The Oura ring uses measures such as light exposure, actigraphy, and estimates of someone's circadian rhythm to determine their bed- and waketime and only measures sleep episodes during the night[10]. This means that daytime naps are not reflected in the total sleep time.

#### Supplementary Note 3 – Graph-regularised SVD

The goal of the decomposition or factorisation of a matrix is often to use the decomposition to find a lower-dimensional approximation to the full matrix. In the context of this study, we aimed to employ such a factorisation to reduce a set of typing modalities to a single measure of typing activity. A well-known factorisation method is singular value decomposition (SVD):

$$X = U\Sigma V^{\top}, \quad (1)$$

where $X$ is an $m \times n$ matrix, $U$ is an $m \times m$ orthogonal matrix, $\Sigma$ is an $m \times n$ diagonal matrix containing the singular values of $X$, and $V$ is an $n \times n$ orthogonal matrix containing the eigenvectors of $X^{\top}X$[11]. By picking only those vectors in $U$ and $V$ corresponding to the largest singular values, this is equivalent to principal component analysis, thereby reducing the dimensionality of the data.

Graph-regularised matrix decomposition attempts to do this decomposition whilst respecting the geometrical structure of the data[12]. The data geometry is represented as a graph that is encoded in a (symmetric) adjacency matrix $A$. Its entries $a_{ij} \in \{0, 1\}$ give the connection strength between nodes $i$ and $j$. In this work, we restrict $a_{ij}$ to be binary but they could also be continuous such as in the case of a weighted graph. To incorporate $A$ into the decomposition, its corresponding graph Laplacian is calculated as $L = D - A$, where $D$ is a diagonal matrix containing the sums of the columns (or rows) of $A$. We then seek a decomposition of $X$ by solving the following problem[12]:

$$\min_{H,W}\left(\|X - HW\|^2 + \lambda Tr(W^{\top}LW)\right), \quad (2)$$

where $H$ is an $m \times k$ matrix, $W$ is a $k \times n$ matrix, and $\lambda$ is a regularisation parameter controlling the smoothness of the decomposition $HW \approx X$. $k$ determines the rank of the approximation, i.e., it represents the dimensionality that we reduce our data to.

Minimising the objective in Equation (2) starts by computing an SVD[13]:

$$XR^{\top^{-1}} = U\Sigma V^{\top}, \quad (3)$$

where $RR^{\top} = I + \lambda L$ is an LU decomposition, in which $I$ is the identity matrix. We can then approximate the SVD by taking only the $k$ largest singular values and the corresponding vectors in $U$ and $V$:

$$XR^{\top^{-1}} \approx \tilde{U}\tilde{\Sigma}\tilde{V}^{\top}. \quad (4)$$

Consequently, by setting

$$\hat{H} \coloneqq \tilde{U}, \quad (5)$$

$$\hat{W} \coloneqq \tilde{\Sigma}\tilde{V}^{\top}R^{-1}, \quad (6)$$

the objective in Equation (2) will be minimised[14].

In this study, the data matrix $X$ contains $m = 4$ typing features derived from a participant's keyboard usage, aggregated into hourly bins. These bins form the nodes of the time graph and are connected to 1) their preceding and subsequent neighbours and 2) the corresponding hours of the preceding and subsequent day. In the case of missing data, we simply remove nodes corresponding to unrecorded days from the graph. This leads to disjoint graphs but poses no problem for the analysis. Since every hour is a node,

we have $n = 24 \cdot d$ nodes, where $d$ is the number of days we have recorded for the participant. Our aim is to reduce the $4 \times 24d$ matrix of typing features $X$ to a $1 \times 24d$ vector which gives us a general measure of typing activity for every hour of every day. This can easily be done by setting $k = 1$ and taking the right-hand factor of the graph-regularised decomposition, $\widehat{W}$, as our activity vector.
The left factor $\widetilde{U}$ forms a basis transformation that can be applied to new data $X^*$ and the associated Laplacian $L^*$. This allows us to 'train' the decomposition on a subset of a participant's data and then apply or 'test' it on the remainder. First we rotate the test data into the same space as $\tilde{\Sigma}\widetilde{V}^\top$:

$$B = \widetilde{U}^\top X^* {R^*}^{\top^{-1}}, \qquad (7)$$

where $R^* {R^*}^\top = I + \lambda L^*$. We then set, analogously to how we constructed $\widehat{W}$:

$$\widehat{W}^* \coloneqq B {R^*}^{-1}. \qquad (8)$$

Since $\widetilde{U}$ only contains four values in our case, it does not require much data to train. We therefore decided to use 10% of a participant's data for training. The remaining 90% was used to predict the number of hours slept, with the exclusion of the first three days after the training period to prevent information from the training set leaking into the test set through temporal autocorrelation. (For the GRSVD of author AL's data, we instead used a 5/95 train-test split due to the large number of available data. In addition, we trained on the tail (the last 5%) rather than the head of that data set.) Participants that did not have enough days of data to make this train-test split (i.e., less than 10 days of data) were excluded from the analysis.
The first component of every GRSVD computed in this study was unipolar, but sometimes negative. Since the component sign is arbitrary, we multiplied such components with -1 to ensure uniformity (positivity) across participants.

#### Supplementary Note 4 – Sleep duration prediction

To predict sleep, the GRSVD scores were binarised into two categories: All values below some fixed threshold (1.0 in the main text) were classified as sleep, and the rest was classified as wakeful. This threshold was determined from visual inspection of the SVD histograms and positioned to be in-between the two peaks of the bimodal distribution. The results of alternative, data-driven thresholding methods are given in Supplementary Figure 15 and Supplementary Figure 16. After classification, the number of hours classified as sleeping were counted to arrive at a prediction for sleep duration. For validation, we used nightly aggregates of the total sleep time as measured by the smart ring and predicted them with our own sleep duration estimates using linear mixed-effects models.

The mixed-effects models included a random intercept $b_{0i}$ and a random slope for typing-derived sleep estimates $b_{1i}$ per participant $i$:

$$y_{ij} = \beta_0 + b_{0i} + \beta_1 x_{ij} + b_{1i} x_{ij} + \epsilon_{ij}, \tag{9}$$

where $y_{ij}$, $x_{ij}$, and $\epsilon_{ij}$ respectively represent the ring-derived total sleep time, typing-derived sleep estimate, and error for participant $i$ and day $j$, while $\beta_0$ and $\beta_1$ give, respectively, the grand mean and fixed effect of our estimate across participants. BiAffect-derived sleep duration was standardised (M = 0, SD = 1) across all observations. Since it is uncommon to test mixed-effects models out of sample, we did not do a train-test split here; whilst several approaches could be used for this, for example witholding a small amount of calibration data to learn the random effects, we consider this to be outside the scope of the current manuscript. Model assumptions were checked using visual inspection of the QQ plots of the residuals and random effects. No gross violations of these assumptions were detected.

#### Supplementary Note 5 – Phase extraction

We converted every entry of author AL's GRSVD matrix to polar coordinates $(r, \theta)$, where the radius $r$ corresponds to the entry's SVD value and the angle $\theta = \frac{h}{24} \cdot 2\pi$ corresponds to the hour of day $h$ rescaled to $[0, 2\pi)$. We then calculated a weighted version of the circular mean for every row (day) of the SVD matrix:

$$\bar{\rho} = \frac{1}{N}\sum_{n=1}^{N} r_n \cdot e^{i\theta_n} = \frac{1}{N}\sum_{n=1}^{N} r_n(cos\,\theta_n + i\,sin\,\theta_n)\,, \qquad (10)$$

where $N = 24$. The angle of this circular mean can then be calculated by:

$$\bar{\theta} = atan2\left(\frac{1}{N}\sum_{n=1}^{N} r_n\,sin\,\theta_n\,, \frac{1}{N}\sum_{n=1}^{N} r_n\,cos\,\theta_n\right). \qquad (11)$$

Similarly, for its radius:

$$\bar{r} = \sqrt{\left(\frac{1}{N}\sum_{n=1}^{N} r_n\,cos\,\theta_n\right)^2 + \left(\frac{1}{N}\sum_{n=1}^{N} r_n\,sin\,\theta_n\right)^2}\,. \qquad (12)$$

### Supplementary Note 6 – CLEAR-3 sample

***Supplementary Table 1***

*Demographics overview. Note that participants could have had multiple diagnoses. SD = Standard deviation. PM = Past month.*

| Variable | Self-report | Oura ring | Phase analysis |
|---|---|---|---|
| N | 69 | 38 | 33 |
| Controls (%) | 0 (0) | 0 (0) | 8 (24.24) |
| Mean age (SD) | 26.75 (4.66) | 26.92 (4.19) | 27 (4.5) |
| Gender (%) | | | |
| Female | 64 (92.75) | 34 (89.47) | 31 (93.94) |
| Nonbinary | 4 (5.8) | 3 (7.89) | 0 (0) |
| Other | 1 (1.45) | 1 (2.63) | 0 (0) |
| Unknown or not reported | 0 (0) | 0 (0) | 2 (6.06) |
| House income (%) | | | |
| less than $15,000 | 5 (8.06) | 4 (12.12) | 3 (9.68) |
| $15,000 - $19,999 | 0 (0) | 0 (0) | 0 (0) |
| $20,000 - $24,999 | 3 (4.84) | 3 (9.09) | 1 (3.23) |
| $25,000 - $29,999 | 4 (6.45) | 3 (9.09) | 0 (0) |
| $30,000 - $34,999 | 2 (3.23) | 2 (6.06) | 1 (3.23) |
| $35,000 - $39,999 | 4 (6.45) | 3 (9.09) | 2 (6.45) |
| $40,000 - $49,999 | 5 (8.06) | 1 (3.03) | 1 (3.23) |
| $50,000 - $79,999 | 16 (25.81) | 9 (27.27) | 10 (32.26) |
| $80,000 - $99,999 | 9 (14.52) | 4 (12.12) | 3 (9.68) |
| $100,000 or above | 14 (22.58) | 4 (12.12) | 7 (22.58) |
| Unknown or not reported | 0 (0) | 0 (0) | 3 (9.68) |
| Baseline diagnosis (%) | | | |
| Attention-Deficit/Hyperactivity Disorder | 14 (20.9) | 8 (22.22) | 5 (20.83) |
| Agoraphobia (Past 6 Months) | 5 (7.46) | 2 (5.56) | 1 (4.17) |
| Binge Eating Disorder (Past 3 Months) | 3 (4.48) | 3 (8.33) | 2 (8.33) |
| Borderline Personality Disorder | 4 (5.97) | 1 (2.78) | 1 (4.17) |
| Generalised Anxiety Disorder (Past 6 Months) | 30 (44.78) | 16 (44.44) | 7 (29.17) |
| Major Depressive Disorder (PM) | 38 (56.72) | 21 (58.33) | 12 (50) |
| Obsessive Compulsive Disorder (PM) | 5 (7.46) | 3 (8.33) | 3 (12.5) |
| Other Specified Anxiety Disorder | 1 (1.49) | 1 (2.78) | 0 (0) |
| Other Specified Trauma- and Stressor-Related Disorder (PM) | 2 (2.99) | 0 (0) | 0 (0) |
| Persistent Depressive Disorder (Past 2 Years) | 27 (40.3) | 15 (41.67) | 11 (45.83) |
| Posttraumatic Stress Disorder (PM) | 12 (17.91) | 6 (16.67) | 2 (8.33) |
| Panic Disorder (PM) | 4 (5.97) | 2 (5.56) | 0 (0) |
| Specific Phobia (Past 6 Months) | 7 (10.45) | 4 (11.11) | 1 (4.17) |
| Social Anxiety Disorder (Past 6 Months) | 24 (35.82) | 9 (25) | 5 (20.83) |
| Alcohol Use Disorder (Past 12 Months) | 9 (13.43) | 4 (11.11) | 4 (16.67) |
| Cannabis (Past 12 Months) | 6 (8.96) | 3 (8.33) | 2 (8.33) |
| Other/Unknown Substance Use Disorder (Past 12 Months) | 1 (1.49) | 0 (0) | 0 (0) |

| | | | |
|---|---|---|---|
| Sedative-Hypnotic-Anxiolytic (Past 12 Months) | 1 (1.49) | 0 (0) | 0 (0) |
| Bipolar II Disorder (PM) | 3 (4.48) | 2 (5.56) | 0 (0) |
| Other Specified Depressive Disorder (PM) | 1 (1.49) | 0 (0) | 1 (4.17) |
| Other Specified Feeding or Eating Disorder (PM) | 3 (4.48) | 1 (2.78) | 0 (0) |
| Other Specified Obsessive Compulsive and Related Disorder (PM) | 1 (1.49) | 1 (2.78) | 1 (4.17) |

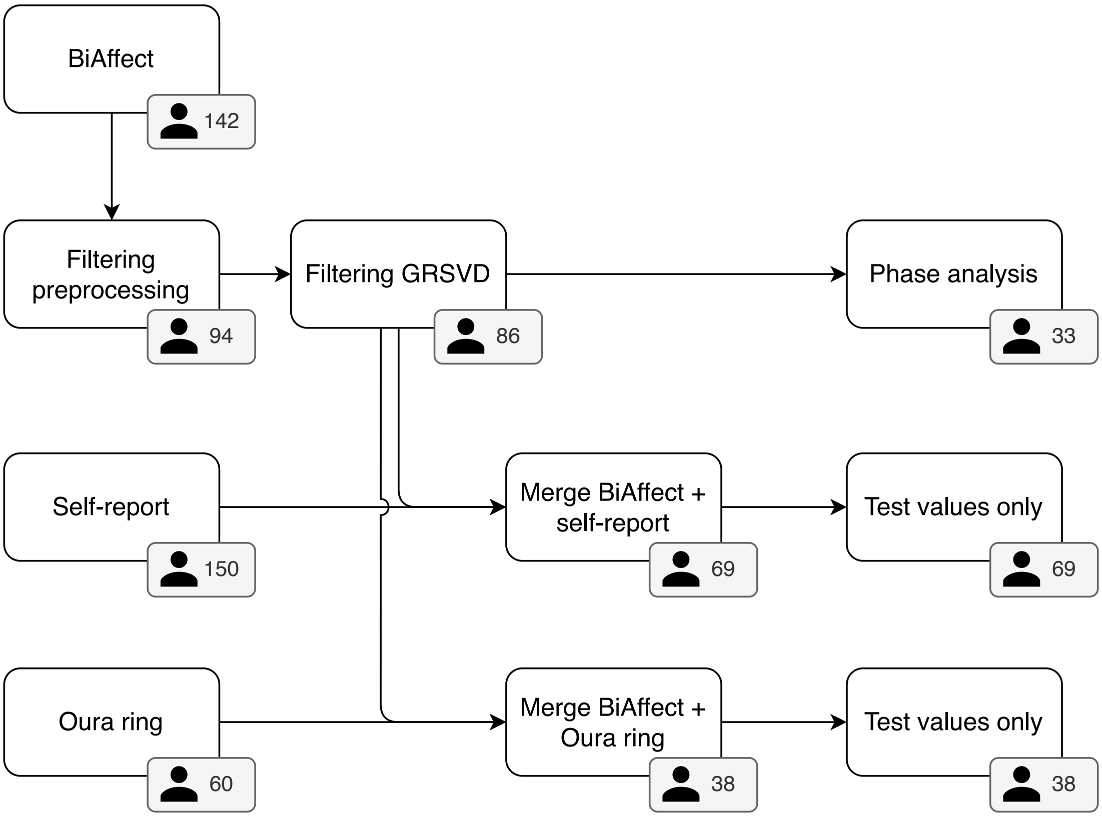

***Supplementary Figure 3: Overview of CLEAR-3 participant exclusion through the processing pipeline.*** *GRSVD = Graph-regularised singular value decomposition.*

The CLEAR-3 trial is a randomised controlled crossover trial that investigated the administration of estradiol (E2) and progesterone (P4) to prevent perimenstrual exacerbation of suicidal ideation due to the natural withdrawal of E2 and P4 in individuals assigned the female sex at birth (AFAB). Participants were recruited via social media. They were 18-45 years of age, had menstrual cycles of 25-35 days, did not take hormonal medication and were in the BMI range 18-34.99. All of them reported past-month suicidal ideation and were in stable outpatient psychiatric treatment. Upon completion of the trial, they received up to US$1250. Participants with any serious long-term nonpsychiatric health condition, a history of hospitalisation for mania or psychosis, or any affective or substance use disorder that would pose a high risk to safe participation in the trial were excluded. All participants provided informed consent.
Supplementary Figure 3 shows how many CLEAR-3 participants were available for the different data sources (BiAffect, self-report, Oura ring) and how their numbers changed

due to filtering and merging of the data streams. Demographics of the final participant groups are given in Supplementary Table 1.

### Supplementary Note 7 – Decomposition overview

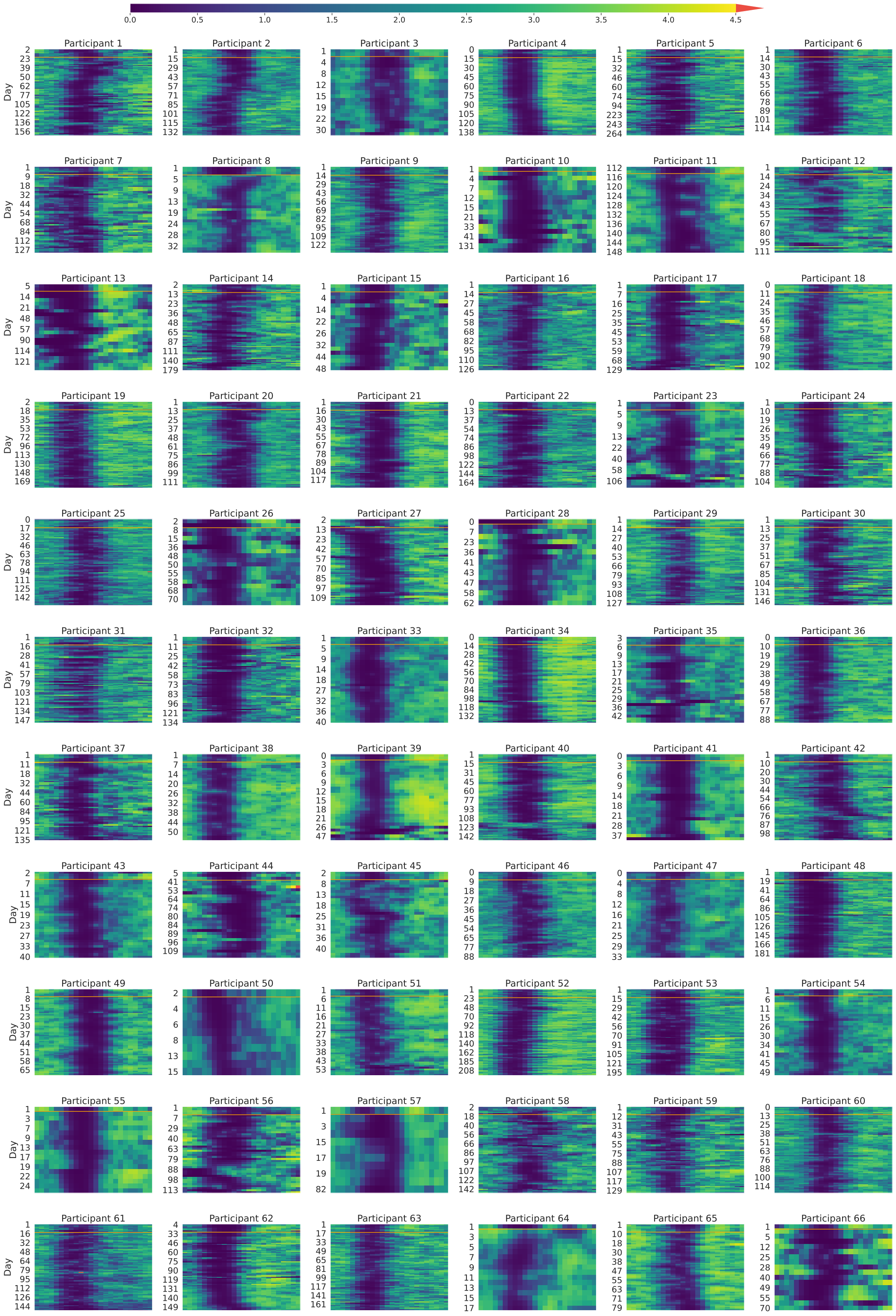

***Supplementary Figure 4: Overview of the graph-regularised SVD results.*** *Colours give a general measure of typing intensity: The brighter the colour, the more typing activity. The horizontal orange line indicates the division between train and test data. Note that the x-axis is in UTC+00:00. Figure continues on next page.*

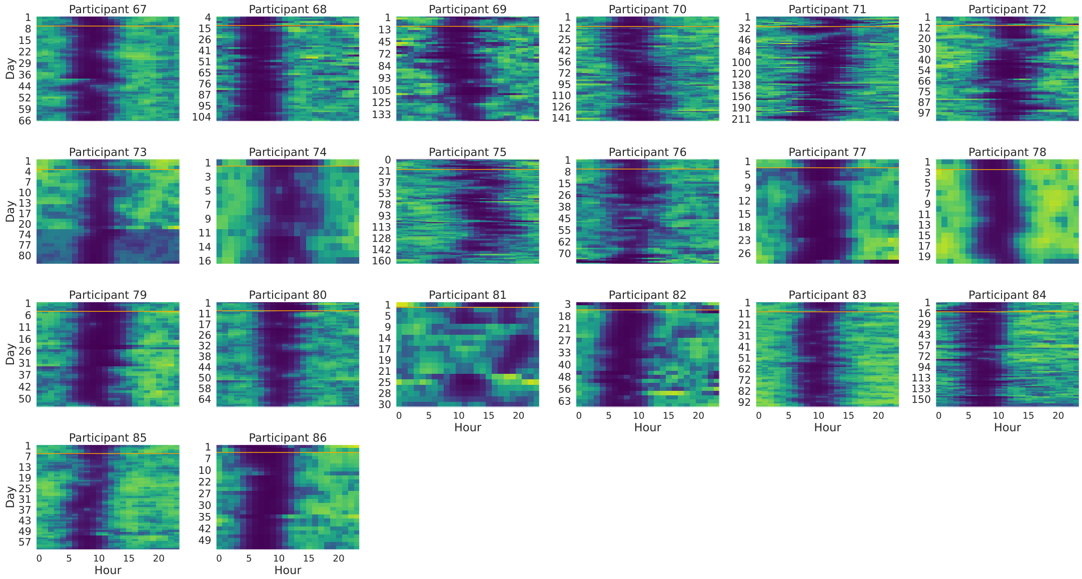

***Supplementary Figure 4 (continued): Overview of the graph-regularised SVD results.***

### Supplementary Note 8 – Statistics phase analysis

***Supplementary Table 2***

*Statistics of the two-sided Welch's t-tests (α = 0.05) employed in the phase analysis in the main text. N denotes the number of transitions for each travel direction. Note that one participant could contribute multiple transitions, and that data for a single transition was not required to be present for all the days surrounding the transition start.*

| Days since transition start | N eastwards | N westwards | Degrees of freedom | Statistic | P-value |
|---|---|---|---|---|---|
| | | | N = 1 transitions | | |
| -3 | 6 | 5 | 5.58 | -1.15 | 0.3 |
| -2 | 6 | 5 | 8.48 | -1.09 | 0.3 |
| -1 | 6 | 5 | 7.35 | 1.49 | 0.18 |
| 0 | 6 | 5 | 8.6 | 3.62 | 0.006 |
| 1 | 6 | 5 | 8.51 | 3.67 | 0.0057 |
| 2 | 6 | 5 | 8.71 | 4.1 | 0.0029 |
| 3 | 6 | 5 | 9 | 4.27 | 0.0021 |
| 4 | 6 | 5 | 9 | 4.54 | 0.0014 |
| 5 | 6 | 5 | 8.99 | 4.54 | 0.0014 |
| 6 | 6 | 5 | 8.99 | 5.6 | 0.00034 |
| 7 | 4 | 5 | 6 | 4.3 | 0.0051 |
| | | | All CLEAR-3 transitions | | |
| -3 | 29 | 44 | 44.07 | -0.32 | 0.75 |
| -2 | 30 | 41 | 67.46 | -0.12 | 0.9 |
| -1 | 32 | 42 | 45.5 | 0.35 | 0.73 |
| 0 | 33 | 50 | 70.7 | 1.52 | 0.13 |
| 1 | 29 | 45 | 41.3 | 2.71 | 0.0097 |
| 2 | 29 | 45 | 45.21 | 3.73 | 0.00053 |
| 3 | 28 | 48 | 35.42 | 2.69 | 0.011 |
| 4 | 30 | 45 | 51.92 | 4.3 | <0.0001 |
| 5 | 28 | 45 | 41.23 | 4.07 | 0.00021 |
| 6 | 27 | 45 | 40.28 | 4.4 | <0.0001 |
| 7 | 27 | 44 | 44.4 | 5.08 | <0.0001 |
| | | | CLEAR-3 transitions with ≥2-hour difference | | |
| -3 | 10 | 4 | 3.44 | 0.6 | 0.59 |
| -2 | 11 | 4 | 3.96 | 0.08 | 0.94 |
| -1 | 12 | 4 | 3.9 | -0.87 | 0.43 |
| 0 | 12 | 4 | 3.31 | 1.7 | 0.18 |
| 1 | 10 | 4 | 6.76 | 3.61 | 0.0091 |
| 2 | 10 | 4 | 4.27 | 3.47 | 0.023 |
| 3 | 8 | 4 | 4.67 | 3.61 | 0.017 |
| 4 | 12 | 4 | 4.17 | 3.4 | 0.026 |
| 5 | 12 | 4 | 4.98 | 3.8 | 0.013 |
| 6 | 10 | 4 | 5.97 | 4.49 | 0.0042 |
| 7 | 11 | 4 | 4.16 | 3.6 | 0.021 |

### Supplementary Note 9 – The Druijff-Van de Woestijne method

To our knowledge, the only other method that utilises smartphone key presses to predict sleep duration has been designed by Druijff-Van de Woestijne and colleagues (DVDW)[15]. In brief, they collected both sleep diaries and keystroke timestamps from a sample of 51 students to predict user-reported metrics such as sleep onset time, wake time, and, most relevant to us, total sleep time (sleep duration). DVDW used mixed-effects models with gender, the number of hours with keystrokes on the previous day, the number of hours with keystrokes on the current day, and a measure called the keystroke-absence period (KAP). The KAP was defined as "the longest time interval without keyboard activity between 19:00 and 15:00 hours (on the next day)"[15]. Their model also included a random intercept per participant.

We implemented the DVDW method and applied it to our own Oura data to be able to compare it with our own graph-regularised SVD (GRSVD) method. To make the comparison fair, we retained the filtering rules applied during initial preprocessing as described above, except that we did not remove days with time zone transitions to reduce the complexity of this sensitivity analysis. Because of the relatively small number of time zone transitions in the CLEAR-3 data, we do not expect this to impact the conclusion. Another difference is that the DVDW method required a different time shift before filtering: Their observation window ranged from 19:00 to 15:00 on the next day, while ours ranged from 12:00 to 12:00 on the next day. In addition, our GSVD method required enough data to make a train-test split. This resulted in 865 observations for 39 participants for the DVDW model, which was more than the 604 observations for the 38 participants in our model. This might bias the comparison slightly in favour of the DVDW model but should not result in large differences.

Estimated coefficients for the DVDW model are shown in Supplementary Table 3. Larger keystroke-absence periods predicted more total sleep time as measured by the Oura ring ($\beta = 0.64$, $p < 0.0001$), whilst more active hours on the current day predicted less total sleep time ($\beta = -0.11$, $p = 0.029$). The latter effect does not correspond to the results found by Druijff-Van de Woestijne et al., who reported no significant effect of hours with keystrokes and besides that a small positive effect of hours with keystrokes on the previous day[15]. The predictions made by the model as well as its total variance breakdown are shown in Supplementary Figure 5. 18.3% of the total variance was explained by the fixed effects, 22.9% by the random intercepts, and 58.8% of the variance remained unexplained. This means that the amount of explained variance (41.2%) is comparable to what is explained by the GSVD model (42.8%). The majority of the point mass is beneath the $y = x$ diagonal, but the distribution does not seem to be as skewed as in the main text.

***Supplementary Table 3***

*Estimated coefficients of the Druijff-Van de Woestijne model. KAP: Keystroke-absence period. SE: Standard error.*

| | **Coefficient** | **SE** | **p-value** |
|---|---|---|---|
| KAP | 0.64 | 0.048 | < 0.0001 |
| # Active hours current day | -0.11 | 0.048 | 0.029 |
| # Active hours previous day | -0.02 | 0.042 | 0.586 |

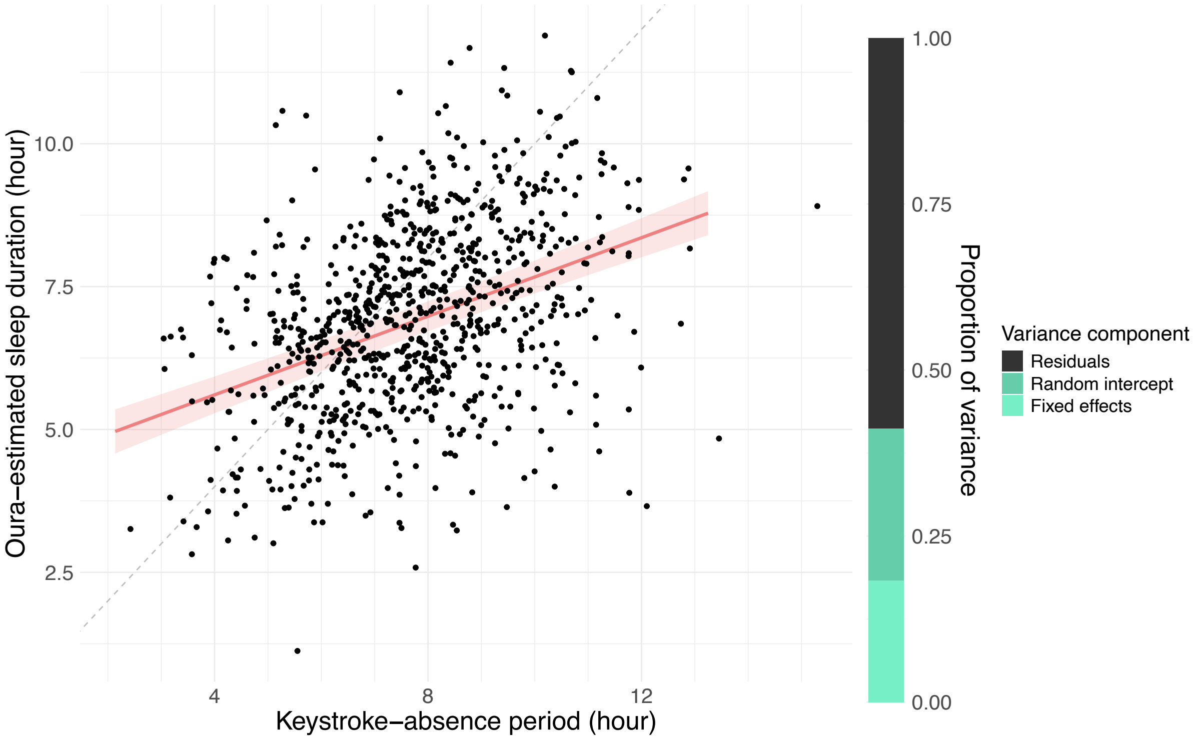

***Supplementary Figure 5: Prediction of the Oura-estimated sleep duration by the Druijff-Van de Woestijne model.***

### Supplementary Note 10 – Varying the data exclusion threshold

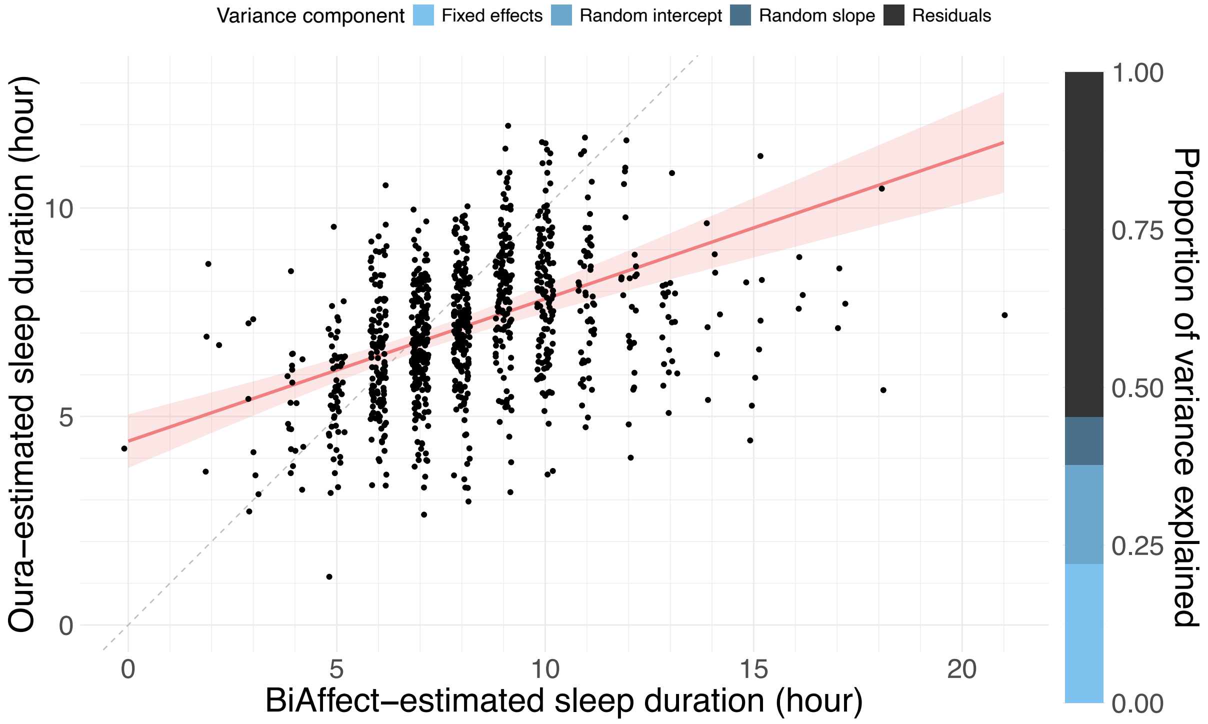

***Supplementary Figure 6: Model predictions and variance decomposition with the four-hour exclusion threshold.***

In the main text, we showed that our method tended to overestimate sleep duration. Part of this overestimation might be due to the participants' underutilisation of their phone, leading to many hours on specific days being marked as inactive while in reality the participant was awake. In general, both a lack of data and poor data quality will cause any machine learning model to perform poorly, so we varied our data exclusion threshold to determine the effect of the data quantity-quality trade-off on our model performance. More specifically, we created two additional models, for one requiring at least four hours and for the other at least twelve hours of each day to contain at least 30 key presses. Note that the model used in the main text required at least eight hours of the day to contain at least 30 key presses.

The four-hour threshold allowed us to include 963 observations from 40 participants. Supplementary Figure 6 shows the associated model predictions and variance decomposition. Centred BiAffect-derived sleep duration is a significant predictor ($\beta = 0.80$, $p < 0.0001$). Supplementary Table 4 contains the numerical explained variance along with the model's RMSE and MAE. Overall, explained variance improves but the RMSE and MAE worsen compared to the eight-hour threshold model of the main text. In addition, with the less restrictive filtering, BiAffect-derived sleep durations display more extreme, implausible values (up to 21 hours).

For the twelve-hour threshold, the number of available observations was reduced to 164 from 22 participants. Supplementary Figure 7 shows the model predictions and variance decomposition. Centred BiAffect-derived sleep duration is a significant predictor ($\beta = 0.66$, $p < 0.0001$). Again, see Supplementary Table 4 for the numerical values and other model metrics. Compared to the eight-hour threshold model, the model displays a

decrease in explained variance but also a decrease in the error metrics RMSE and MAE. BiAffect-derived sleep durations are less extreme, only going up to 10 hours.
Overall, this suggests that using stricter thresholds (including less data) leads to worse explained variance, better error metrics, and fewer BiAffect-derived sleep durations.

***Supplementary Table 4***

*Explained proportions of the total variance and error metrics for models with different data exclusion thresholds. Note that the amount of data is not equal for the different scenarios. Explained variances might not add up to 100% exactly due to rounding.*

| | Four-hour threshold | Eight-hour threshold | Twelve-hour threshold |
|---|---|---|---|
| Fixed effects | 22.0% | 20.7% | 18.5% |
| Random intercept | 15.6% | 17.4% | 16.7% |
| Random slope | 7.7% | 4.7% | 1.1% |
| Residuals | 54.6% | 57.2% | 63.7% |
| RMSE | 1.22 | 1.18 | 1.17 |
| MAE | 0.93 | 0.90 | 0.86 |
| Fixed-effect RMSE | 1.54 | 1.47 | 1.34 |
| Fixed-effect MAE | 1.21 | 1.16 | 1.04 |

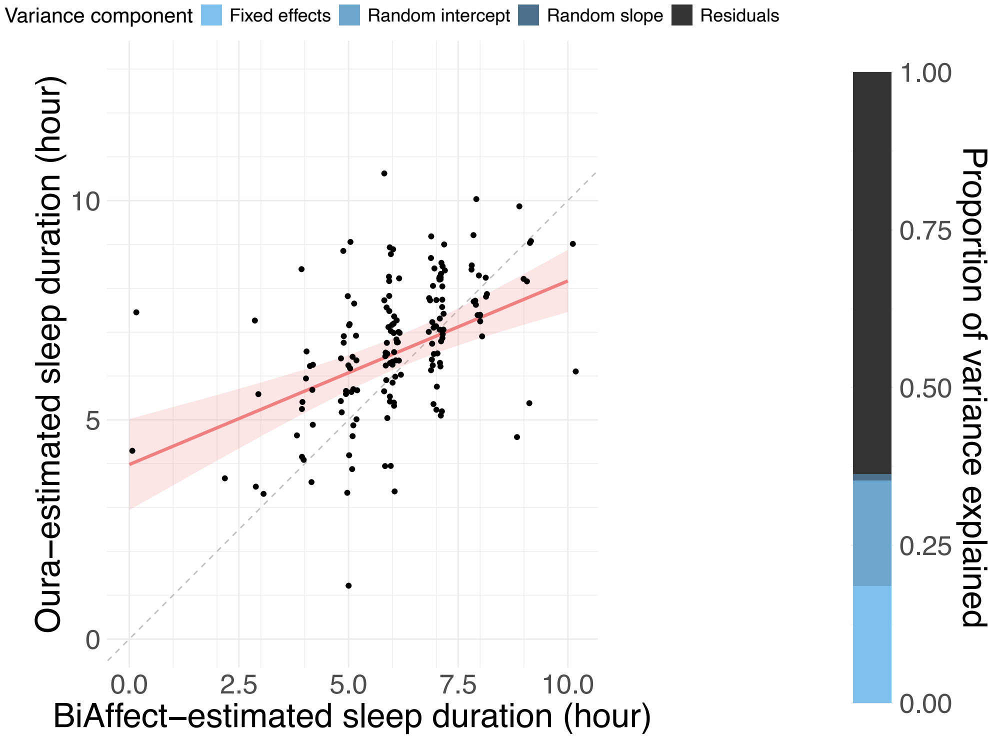

***Supplementary Figure 7: Model predictions and variance decomposition with the twelve-hour exclusion threshold.***

### Supplementary note 11 – Varying the regularisation parameter

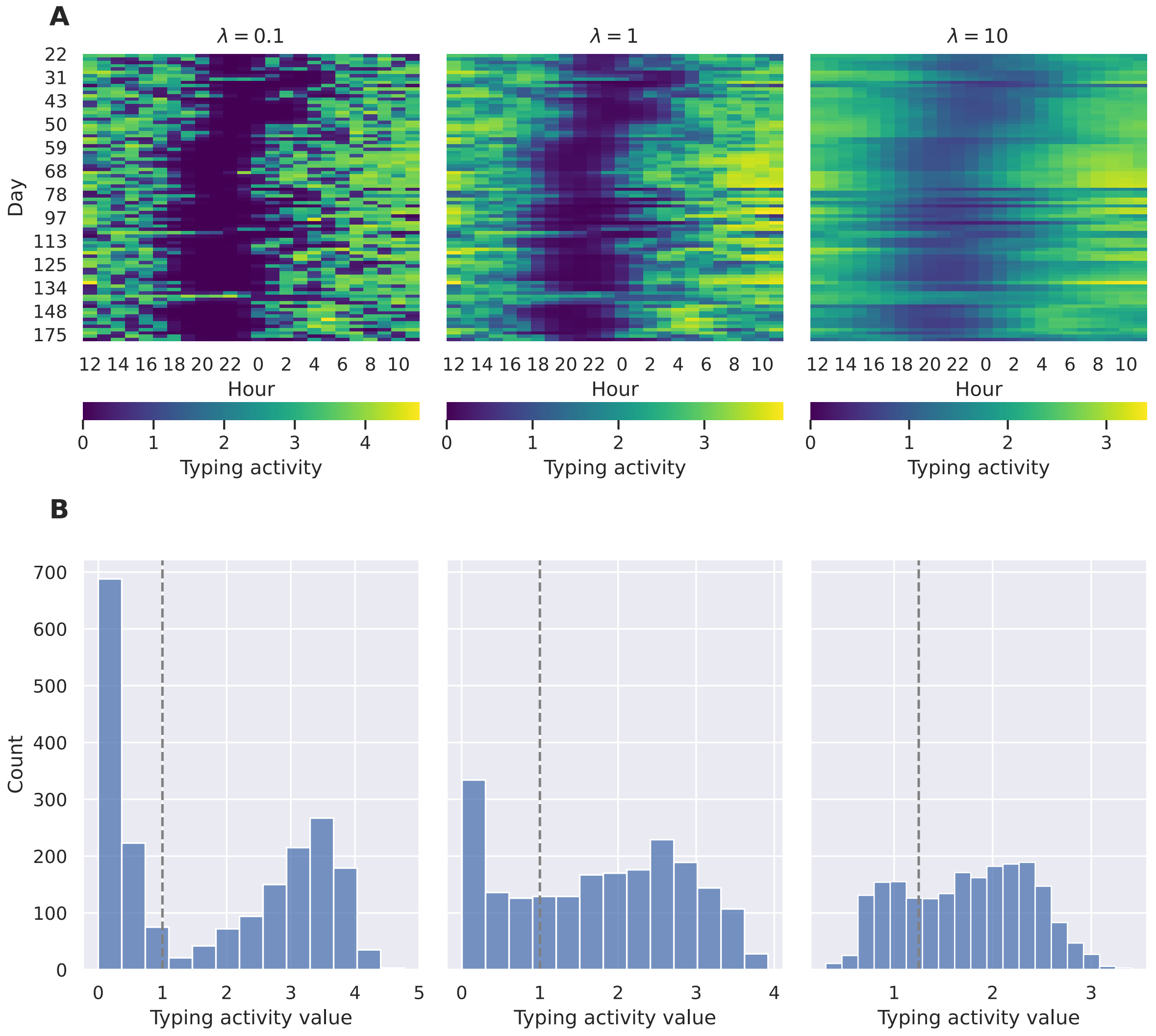

***Supplementary Figure 8: Results of the graph-regularised SVD for a single CLEAR-3 participant using different levels of regularisation. A*** *GRSVD matrices for low regularisation (λ = 0.1), medium regularisation (λ = 1, used in the main text) and high regularisation (λ = 10).* ***B*** *Histograms of the values shown in the matrices above. Note that the range of values in the matrices is not equal across regularisation levels.*

The amount of graph regularisation applied in our SVD method is dependent on the regularisation parameter $\lambda$ (see Equation (2)). To assess its influence on the decomposition and subsequent sleep predictions, we reran our algorithms with $\lambda = 0.1$ and $\lambda = 10$. Supplementary Figure 8A shows the resulting decompositions for a single participant. As expected, increasing $\lambda$ leads to smoother decompositions. Especially at higher levels, discontinuities in the underlying graph become visible in the decomposition as horizontal lines that interrupt the gradient of colours. This is what the graph-regularised SVD was designed to do: Connected nodes should have similar values, disconnected ones are free to vary.

Supplementary Figure 8B shows how the distribution of GRSVD scores (typing activity values) changes with higher values of $\lambda$. The bimodal pattern that is evident at lower levels of regularisation becomes less clear as the amount of regularisation increases. At high regularisation levels, there is more variability in where the dip between the two peaks appears across participants (not shown here), and sometimes the bimodal distribution disappears altogether. This is a natural consequence of the increased amount of smoothing that comes with higher values of $\lambda$: Areas in the data matrix with no data will get mixed with those areas that do contain data, yielding a homogeneous blob when the smoothing is pushed to the extreme. Fortunately, our cases here are not that extreme, so we chose a binarisation threshold of 1.0 for $\lambda = 0.1$ and 1.25 for $\lambda = 10$ based on visual inspection.

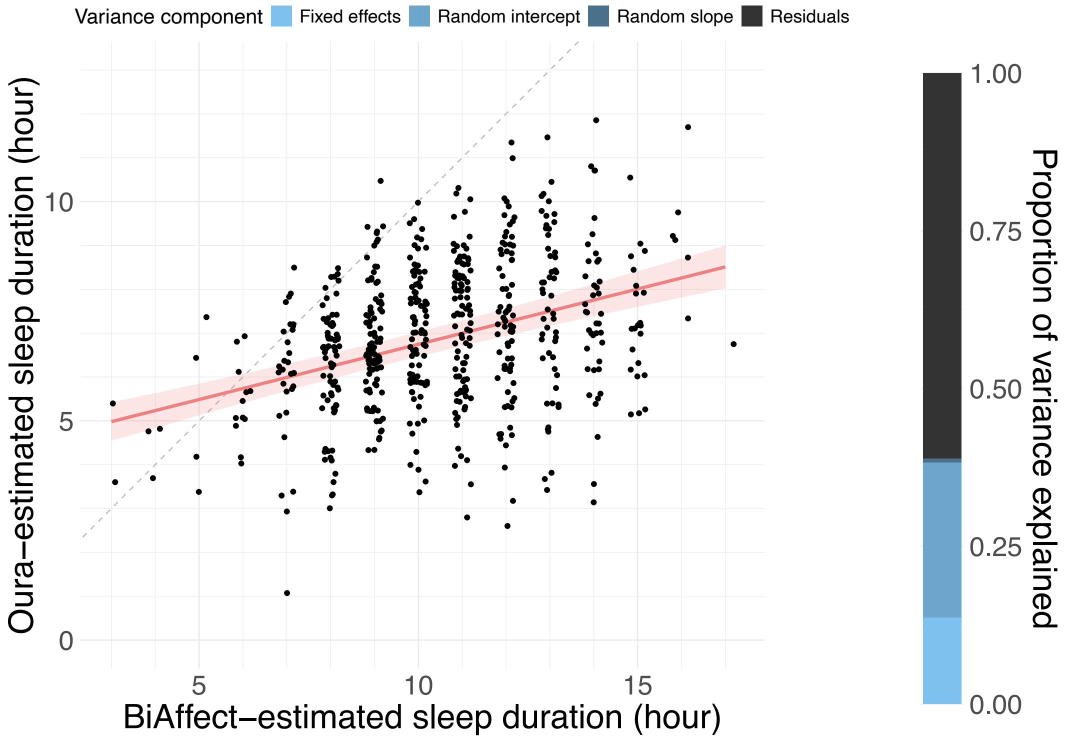

***Supplementary Figure 9: Model predictions for the $\lambda = 0.1$ data.***

Model predictions for the $\lambda = 0.1$ data are given in Supplementary Figure 9. Although the BiAffect-derived estimate is a significant predictor of sleep duration (β = 0.60, p < 0.0001), the overestimation of sleep duration has become much more severe relative to the main text. Explained variance and error metrics have also become slightly worse (see Supplementary Table 5).

***Supplementary Table 5***

*Explained proportions of the total variance and error metrics for models with different values for the regularization parameter $\lambda$. Contrary to Supplementary Table 4, the amount of data is the same for all scenarios.*

| | $\lambda = 0.1$ | $\lambda = 1$ | $\lambda = 10$ |
|---|---|---|---|
| Fixed effects | 13.7% | 20.7% | 12.9% |
| Random intercept | 24.5% | 17.4% | 20.2% |
| Random slope | 0.7% | 4.7% | 2.5% |
| Residuals | 61.0% | 57.2% | 64.4% |
| RMSE | 1.22 | 1.18 | 1.28 |
| MAE | 0.93 | 0.90 | 0.96 |
| Fixed-effect RMSE | 1.55 | 1.47 | 1.57 |
| Fixed-effect MAE | 1.24 | 1.16 | 1.26 |

The model predictions for $\lambda = 10$ are shown in Supplementary Figure 10. The predictions seem to be more balanced in terms of under- and overestimation, although they are leaning more towards the latter. The prediction is still significant (β = 0.59, p < 0.0001). Again, explained variance and error metrics are worse than for the main text model (see Supplementary Table 5).

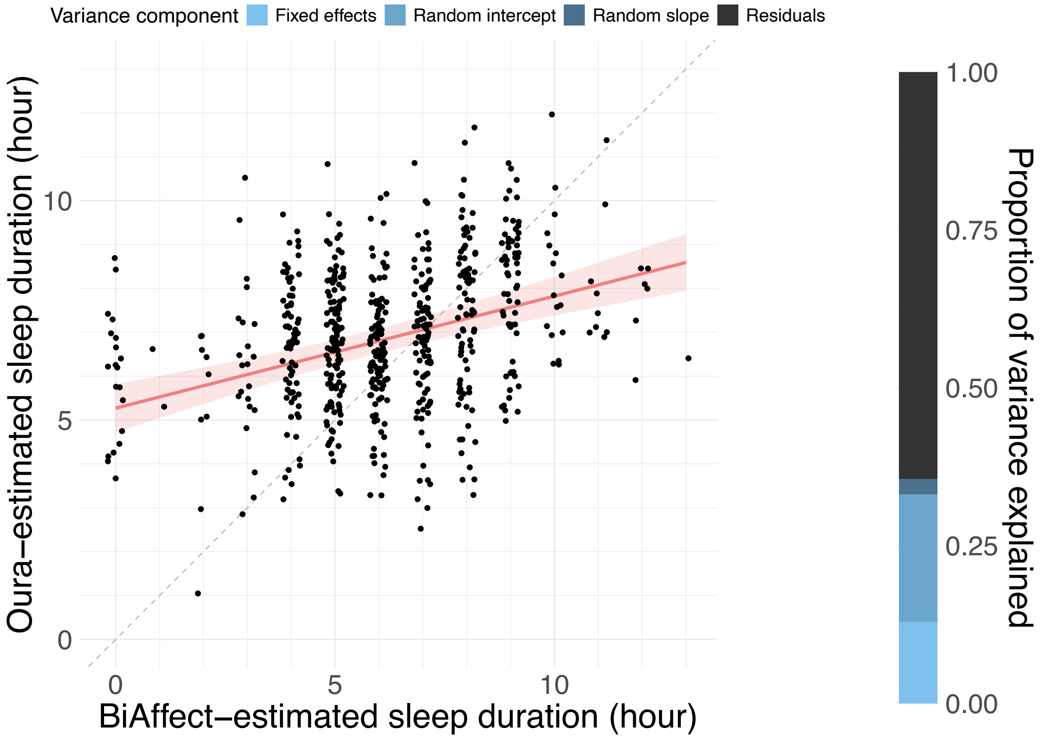

***Supplementary Figure 10: Model predictions for the $\lambda = 10$ data.***

#### Supplementary Note 12 – Predictions using self-report data

Besides Oura ring data, the CLEAR-3 study also collected daily self-report data on participants' sleep duration. Participants could indicate their sleep duration in hours using a nine-point Likert scale ranging from "2 or less hours of sleep" to "10 or more hours of sleep".

We used the self-report data as a dependent variable being predicted from the BiAffect-derived sleep duration estimates using a mixed-effects model with random intercepts and slopes for participants (Supplementary Figure 11). BiAffect-derived sleep duration was centred and scaled to unit variance. Our sleep duration estimates significantly predicted self-reported sleep duration ($\beta = 0.50$, $p < 0.0001$), explaining 11.3% of the total variance, with random intercepts and slopes respectively explaining 25.6% and 4.1% of the variance. 59.1% therefore consisted of residual variance. The model displayed an RMSE of 1.10 and an MAE of 0.83. RMSE and MAE incorporating only the model's fixed effects were 1.41 and 1.13, respectively.

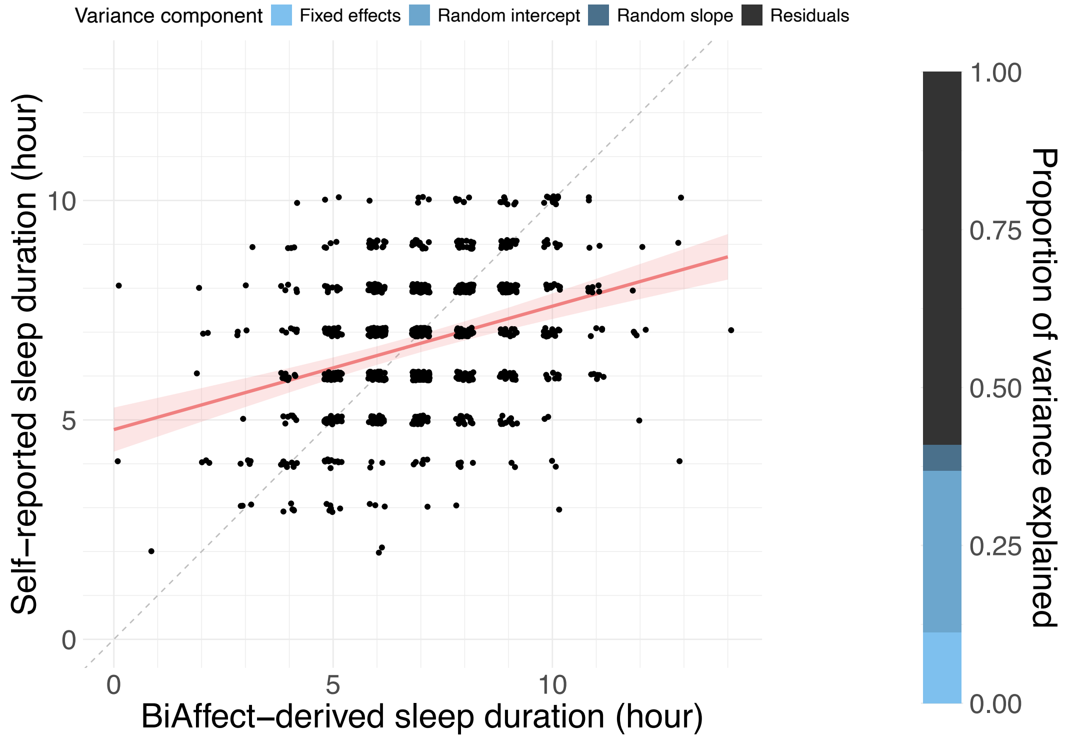

***Supplementary Figure 11: Model predictions of the self-report data with BiAffect-derived sleep estimates as the dependent variable.*** *Jittered data points are plotted over the regression line. The shaded band represents the 95% CI.*

We also found that the self-report data (which was centred and scaled to unit variance) strongly predicts the Oura ring data in a model with participant-level random intercepts and slopes ($\beta = 1.26$, $p < 0.0001$). The variance explained by the fixed effects is 52.6% of the total variance, while the random intercepts and slopes explain 1.6% and 6.0%, respectively (see Supplementary Figure 12). The model displayed an RMSE of 0.93 and an MAE of 0.64; fixed-effect RMSE and MAE were 1.03 and 0.72, respectively. These metrics are better than with typing-derived sleep estimates, but it should be noted that participants were able to view their Oura sleep metrics during the study, which might have biased their self-reports.

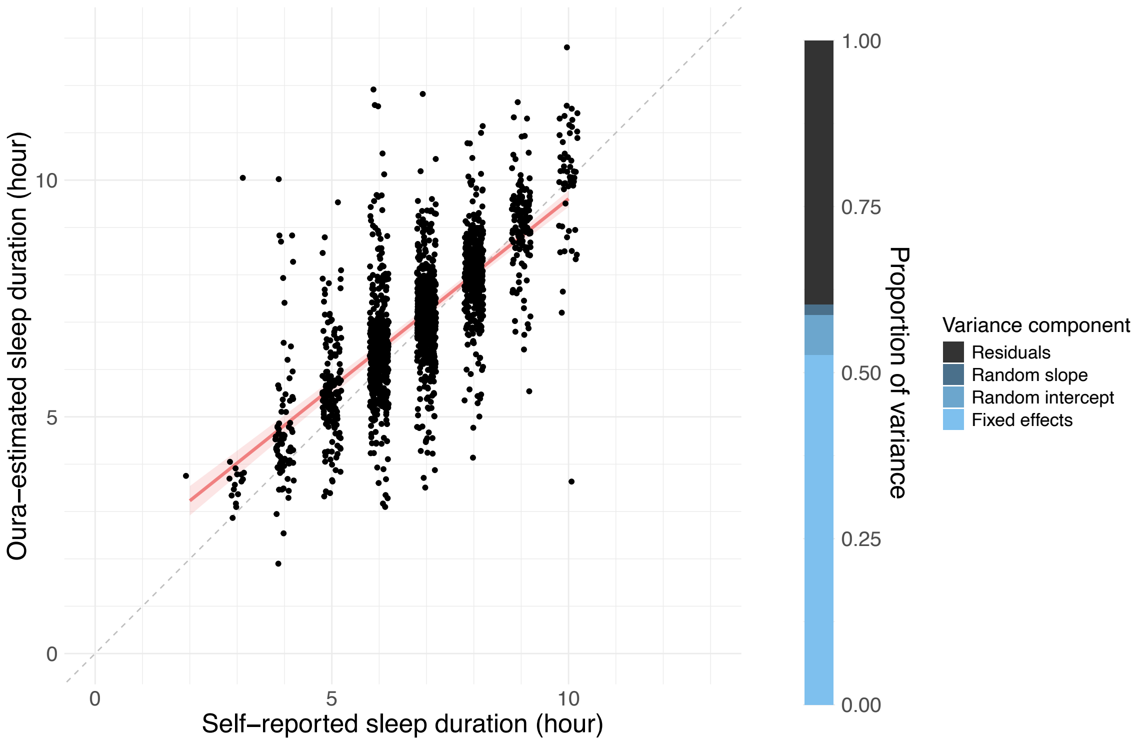

***Supplementary Figure 12: Model predictions of the Oura ring data with self-report as the independent variable.*** *Jittered data points are plotted over the regression line. The shaded band represents the 95% CI.*

### Supplementary Note 13 – Full-rank decomposition

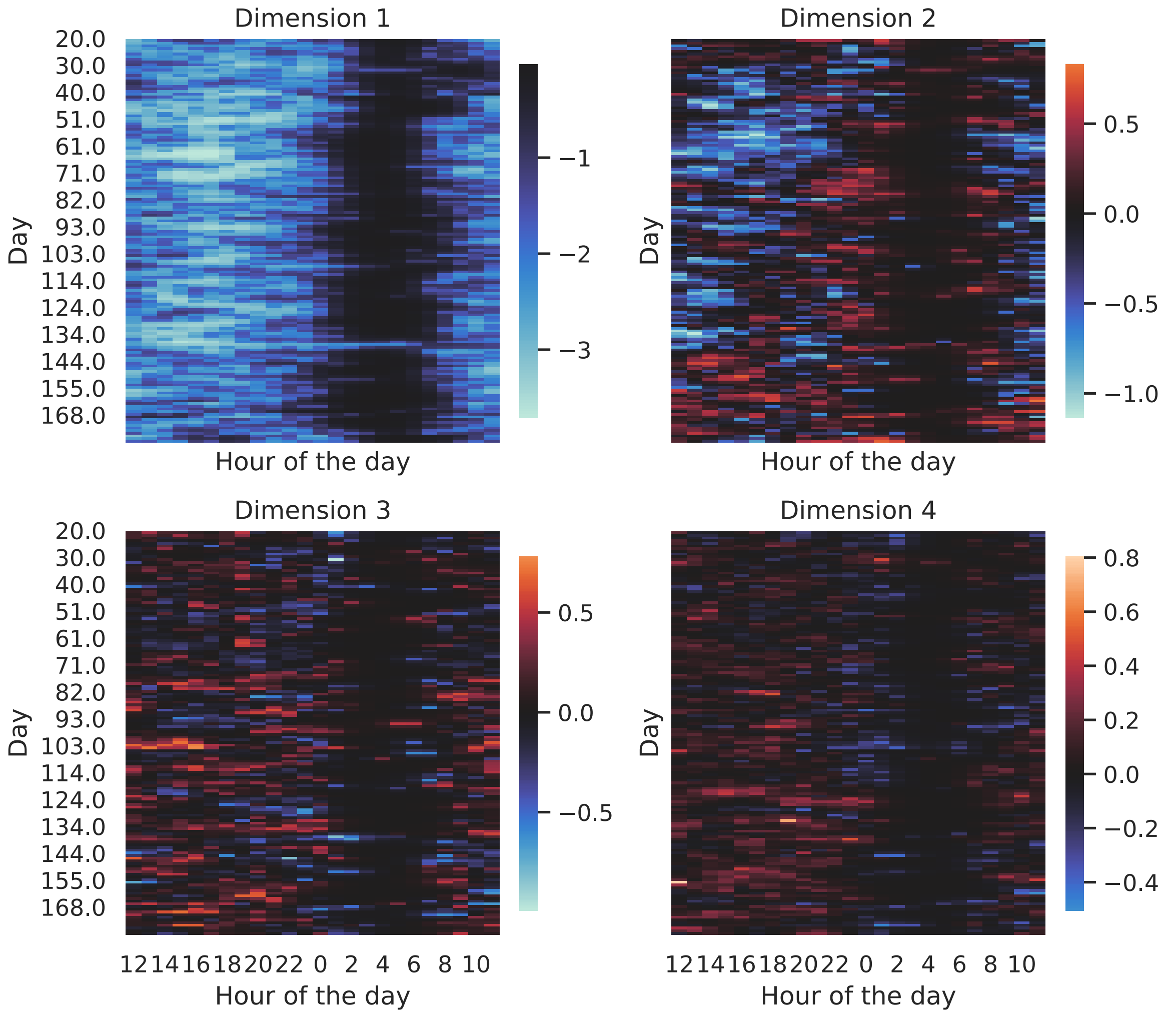

***Supplementary Figure 13: Full-rank decomposition of the test data of a sample participant.***

Supplementary Figure 13 shows a full-rank decomposition of a participant's data matrix. In the main text, we only use the first dimension, in this case multiplied by -1 to ensure positivity (as mentioned above, the sign of the components of an SVD is arbitrary). Whereas the first dimension shows a strong dichotomy between sleep and wakeful periods, the other dimensions do not show such a strong division and mainly seem to be concerned with intraday variations in typing metrics, which makes it unclear how to summarise them to daily measures suitable for sleep prediction.

### Supplementary Note 14 – Circular mean characteristics

The main text discusses the angle of the circular mean of author AL's daily activity profile, which can be interpreted as the phase of her circadian rhythm. In Supplementary Figure 14, we show this phase over a long time span along with its first derivative, visualising the rate of change of the rhythm phase, and the magnitude of the circular mean, which can be interpreted as the strength of the rhythm (larger differences between day- and nighttime activity result in larger magnitudes).

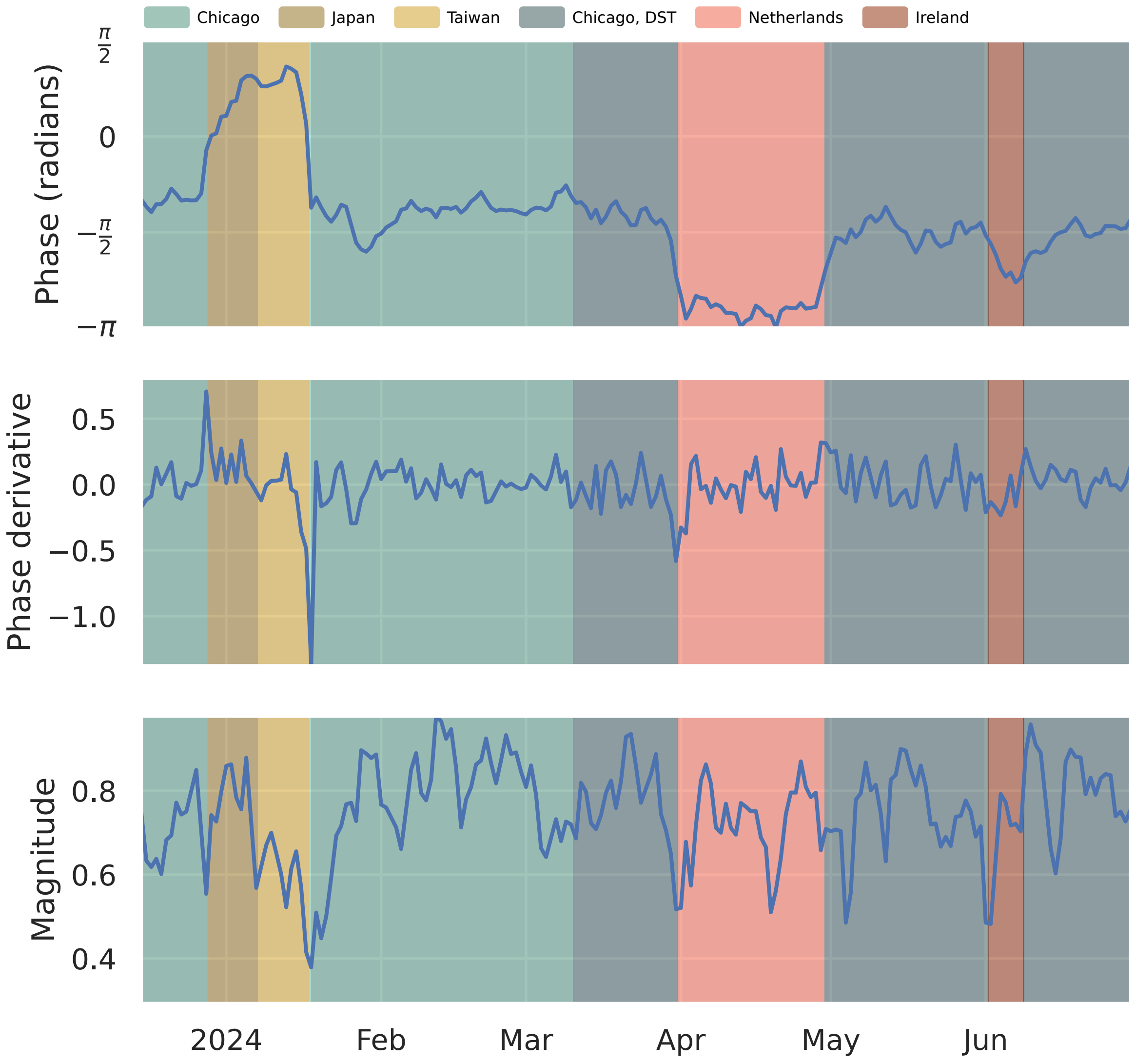

***Supplementary Figure 14: Overview of the circular mean characteristics of AL's trip to Japan.*** *Top: Angle of the circular mean of the daily activity profiles, representing rhythm phase. Same as in Figure 3E in the main text. Middle: The first derivative of the phase, showing its rate of change. Bottom: The magnitude of the circular means. Represents rhythm strength.*

The phase derivative confirms what was already noted in the main text: The phase transition that occurred when AL travelled from Chicago to Japan was gradual, while the transition from Taiwan to Chicago was large and sudden. In addition, the sequence of spikes in the derivative during the gradual Chicago-Japan transition seems to suggest that that transition occurred in discrete steps, but we note that this could be an artifact of the temporal resolution.

The magnitude of the circular means is less smooth than the phase, suggesting that there were many changes in rhythm strength. Some of the largest dips in strength seemed to have occurred around the time zone transitions, especially the Taiwan-Chicago transition. This conforms to what we have seen in Figure 3B and 3C in the main text: For this period, Figure 3B does not show a clear day-night cycle in the SVD matrix, and Figure 3C shows large disturbances in the oscillations of the SVD values. Other peaks and troughs do not line up with time zone transitions, however, and might have come about through other factors affecting AL's circadian rhythm.

#### Supplementary Note 15 – Alternative thresholding methods

To binarise the entries of the GRSVD matrix into 'waking' and 'sleeping', we had to employ a threshold above which entries were marked as 'waking'. In the main text, we chose a fixed threshold of 1.0 for all participants. Here, we explore two alternative, data-driven methods that allow for varying thresholds between participants.

First, we use Otsu's method, an image processing technique used to separate grey-level pixels into two classes[16]. We used the scikit-image package (version 0.20.0) to segment each participant's GRSVD matrix separately. Supplementary Figure 15 shows the predictions of the model built with the resulting sleep durations. Centred BiAffect-derived sleep duration is a significant predictor (β = 0.51, $p < 0.0001$). The model displays a strong tendency to overestimate the true sleep duration, as evidenced by the data points being largely situated under the x=y diagonal (dashed line). 10.0% of the variance was explained by the fixed effects, 23.0% by the random intercepts, and 2.3% by the random slopes, leaving 64.7% unexplained. It showed an RMSE of 1.25 and an MAE of 0.94; fixed-effect RMSE and MAE were 1.59 and 1.26, respectively. These metrics indicate the model is inferior to the one in the main text.

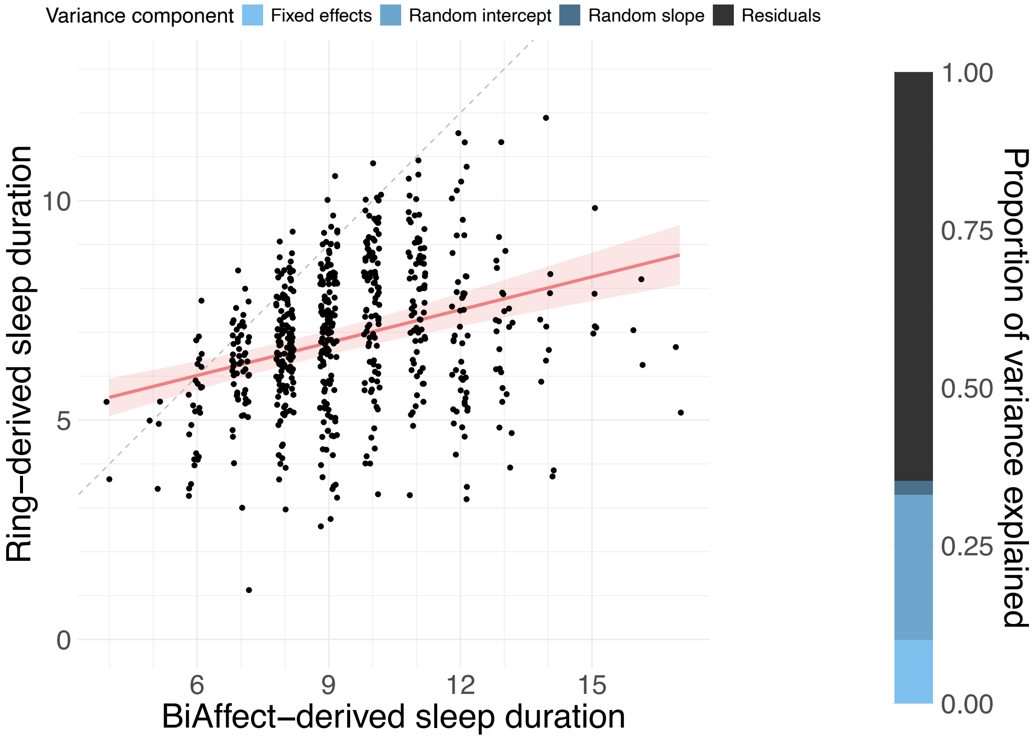

***Supplementary Figure 15: Model predictions with sleep durations derived via Otsu's method.*** *Jittered data points are plotted over the regression line. The shaded band represents the 95% CI.*

Second, we used a Gaussian mixture model with two Gaussians to separate the GRSVD scores of each GRSVD matrix into two classes, taking as threshold the quantile where the densities of the fitted distributions crossed. The mixture was fitted using scikit-learn (version 1.2.2). Model predictions with the derived sleep durations are shown in Supplementary Figure 16. Centred BiAffect-derived sleep duration is a significant predictor (β = 0.61, $p < 0.0001$). Overestimation is much less severe than with Otsu's threshold. 11.8% of the variance is explained by the fixed effects, 36.1% by the random intercepts, 3.9% by the random slopes, and 48.2% is left unexplained. The model shows an RMSE of 1.20 and an MAE of 0.89, with a fixed-effect RMSE and MAE of 1.69 and 1.31, respectively. Both the MAE and total explained variance of this model are better than the respective metrics of the main text model. However, the increase in explained variance is solely due to the random intercepts; the part of the variance explained by the fixed effects has decreased. In addition, the RMSE is worse. In sum, we prefer the main text model (with fixed thresholds) over this model (with Gaussian mixture thresholds).

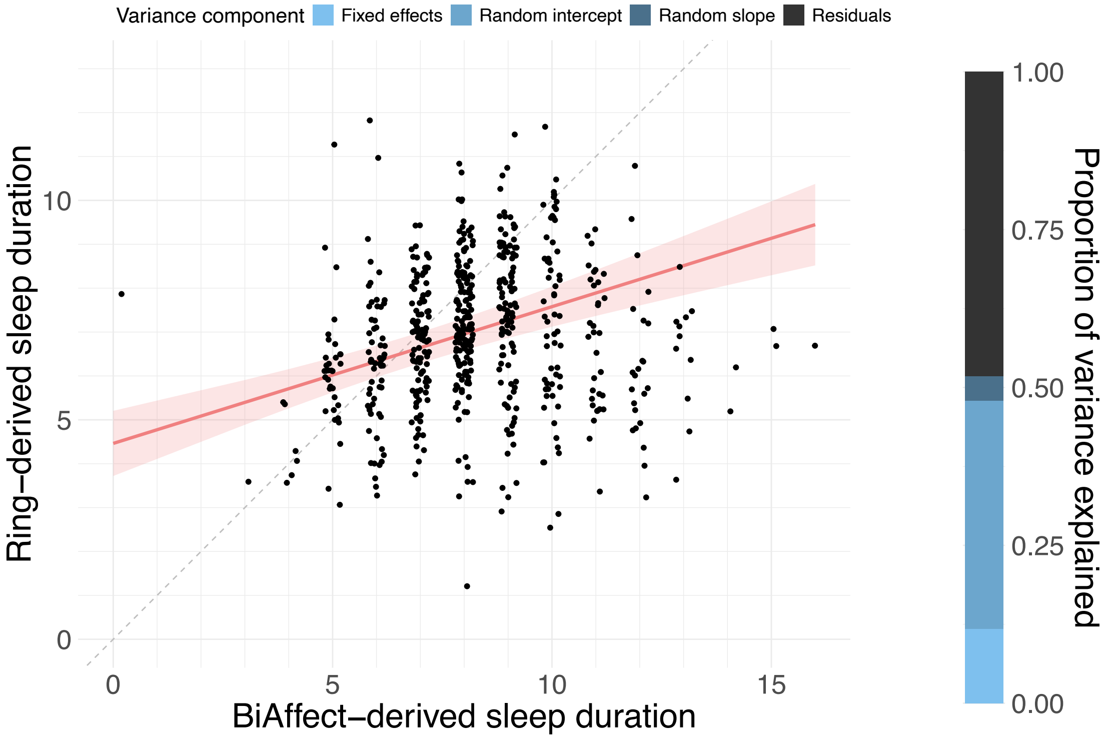

***Supplementary Figure 16: Model predictions with sleep durations derived via a mixture of Gaussians.*** *Jittered data points are plotted over the regression line. The shaded band represents the 95% CI.*

#### Supplementary Note 16 – Predict time in bed

Although we predict sleep duration in the main text, some would argue that what is measured by BiAffect is closer to total recording time (TRT), which is defined as “the total amount of time during which the patient is in bed with recording equipment activated” in the context of polysomnography[17]. In the context of BiAffect, this definition is not without issues, as someone could very well lie in bed while still using their phone. Nevertheless, we extracted time spent in bed (TIB) from the Oura ring data and created a mixed model predicting TIB from the BiAffect-derived sleep durations.

Supplementary Figure 17 shows the model predictions. Centred BiAffect-derived sleep duration is a significant predictor ($\beta = 0.83$, $p < 0.0001$). The correlation between the two variables is slightly better than what we found in the main text (Spearman’s rank correlation $r_s = 0.48$ [$n = 648$, $p < 0.0001$]) and overestimation of TIB is less severe than the overestimation of Oura-derived sleep duration. However, 17.1% of the variance was explained by the fixed effects, 15.4% by the random intercepts, 4.1% by the random effects, and 63.4% was left unexplained. The RMSE was 1.46 and the MAE was 1.09; the fixed-effect RMSE and MAE was 1.75 and 1.35, respectively. All these metrics are slightly worse than what we found in the main text, so we still prefer the latter.

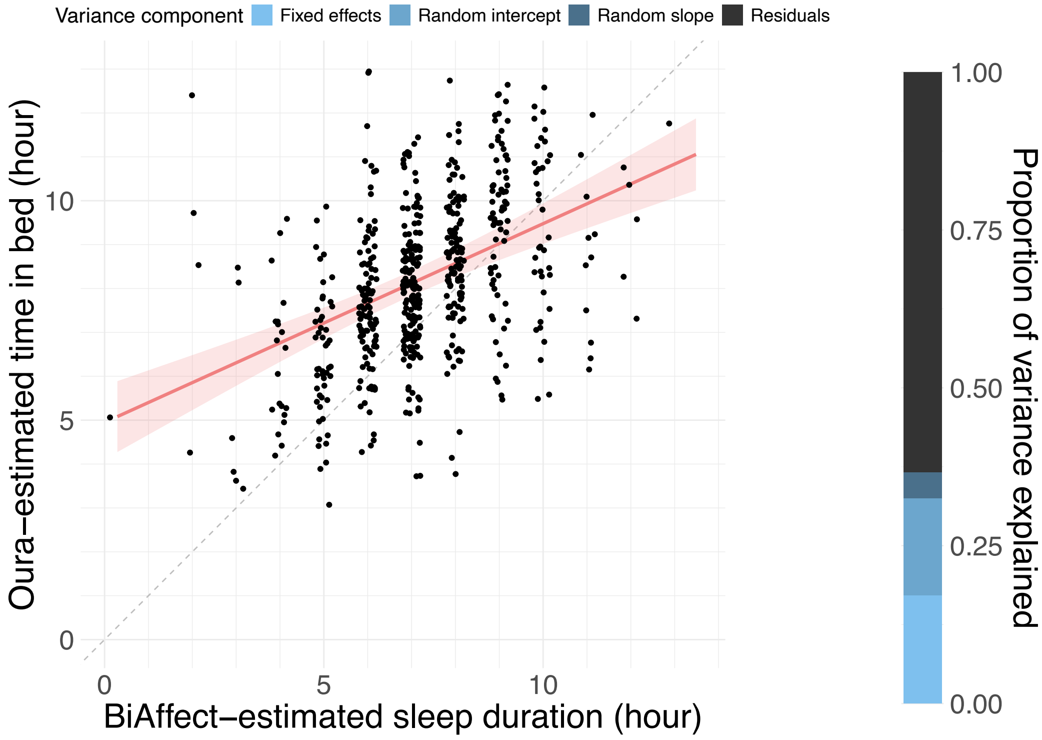

***Supplementary Figure 17: Model predictions of time spent in bed based on BiAffect-derived sleep duration.*** *Jittered data points are plotted over the regression line. The shaded band represents the 95% CI.*

#### Supplementary Note 17 – Add extra modalities

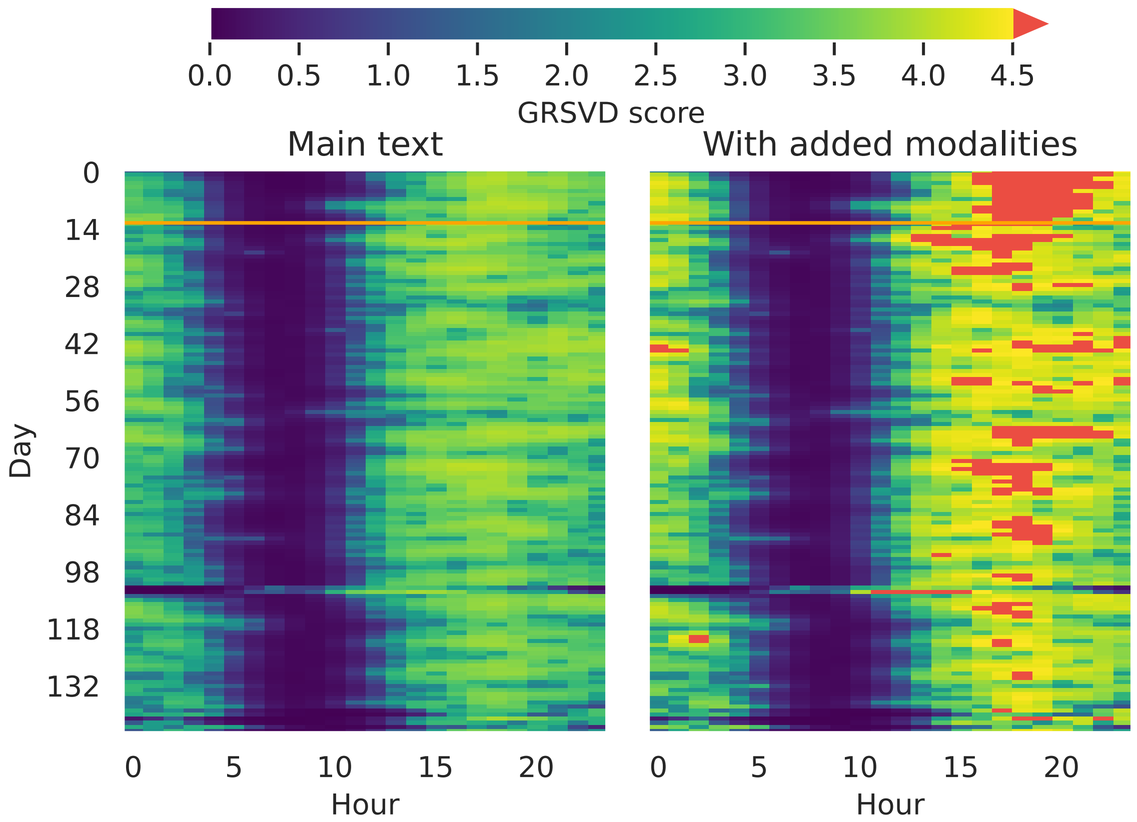

***Supplementary Figure 18: GRSVD matrices of a sample participant, with and without added modalities.*** *Left panel: GRSVD matrix as used in the main text, based on the movement rate, upright rate, number of key presses and typing speed. Right panel: GRSVD matrix based on the aforementioned modalities plus typing speed variability, autocorrect rate, and backspace rate. Colour bar corresponds to the one used in Supplementary Figure 4. Red indicates GRSVD scores over 4.5. GRSVD: Graph-regularised singular value decomposition.*

Since the GRSVD is a dimensionality reduction algorithm, extra modalities are easily added. Here we show the change in GRSVD scores and subsequent sleep duration predictions when adding typing speed variability (mean absolute deviation of the inter-key delay), backspace rate, and autocorrect rate (Supplementary Figure 18). The patterns of high and low GRSVD scores look comparable, but the range of scores seems to be larger when adding modalities. The comparable patterns were to be expected, as the added modalities are recorded at the same time as all the other modalities, namely when a participant starts typing, and will therefore record high and low activity during similar periods. For more profound changes, modalities from other devices would have to be added (e.g., actigraphy). We consider collecting and processing such data to be outside the scope of the current study.

To accommodate the increased range of GRSVD scores, we used a threshold of 1.5 instead of 1.0 for binarising the GRSVD matrix. Model predictions for sleep duration based on the added modalities are shown in Supplementary Figure 19. Centred BiAffect-derived sleep duration was a significant predictor of Oura-derived sleep duration (β = 0.71, $p < 0.0001$). 19.4% of the variance was explained by the fixed effects, 20.2% by the random intercepts, and 3.5% by the random slopes, leaving 56.9% unexplained. The RMSE was 1.18, the MAE was 0.89; fixed-effect RMSE and MAE was 1.49 and 1.18, respectively. Overall, this model was on par with the main text model.

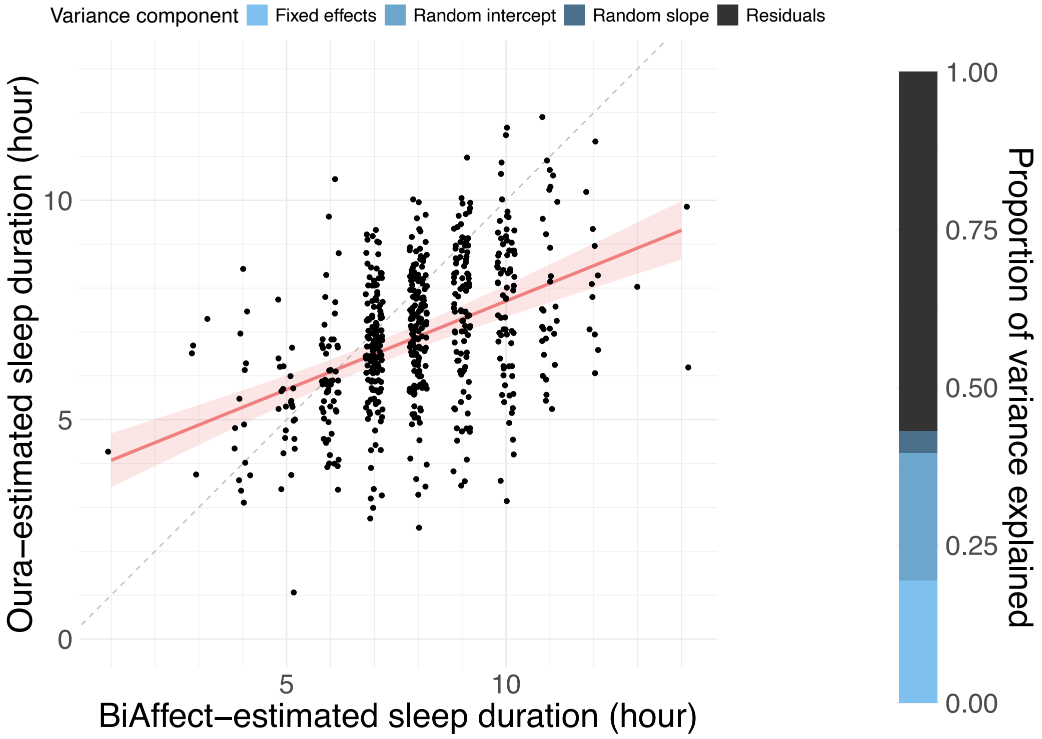

***Supplementary Figure 19: Model predictions using additional typing modalities.*** *Jittered data points are plotted over the regression line. The shaded band represents the 95% CI.*

#### Supplementary Note 18 – Clip BiAffect-derived sleep durations

We optimised our method to provide BiAffect-derived sleep durations in a reasonable range, but it was unfortunately not possible to prevent all implausible values (e.g., 0 hours of sleep). As a sensitivity analysis, we decided to clip extreme values to a range of 1-13 hours, where the range limits were based on the extreme values found in the Oura data. Model predictions for the clipped data can be found in Supplementary Figure 20. Clipped and subsequently centred BiAffect-derived sleep durations significantly predicted Oura-derived sleep duration ($\beta = 0.73$, $p < 0.0001$). 20.6% of the variance was explained by the fixed effects, 17.4% by the random intercepts, 4.8% by the random slopes, and 57.2% of the variance was left unexplained. The RMSE was 1.18 and the MAE was 0.90; fixed-effect RMSE and MAE was 1.47 and 1.16, respectively. All in all, this model is on par with the main text model, and clipping the extreme outliers does not seem to have a large effect on the model performance. Given that here only one data point fell outside the clipping range, we do not find this surprising.

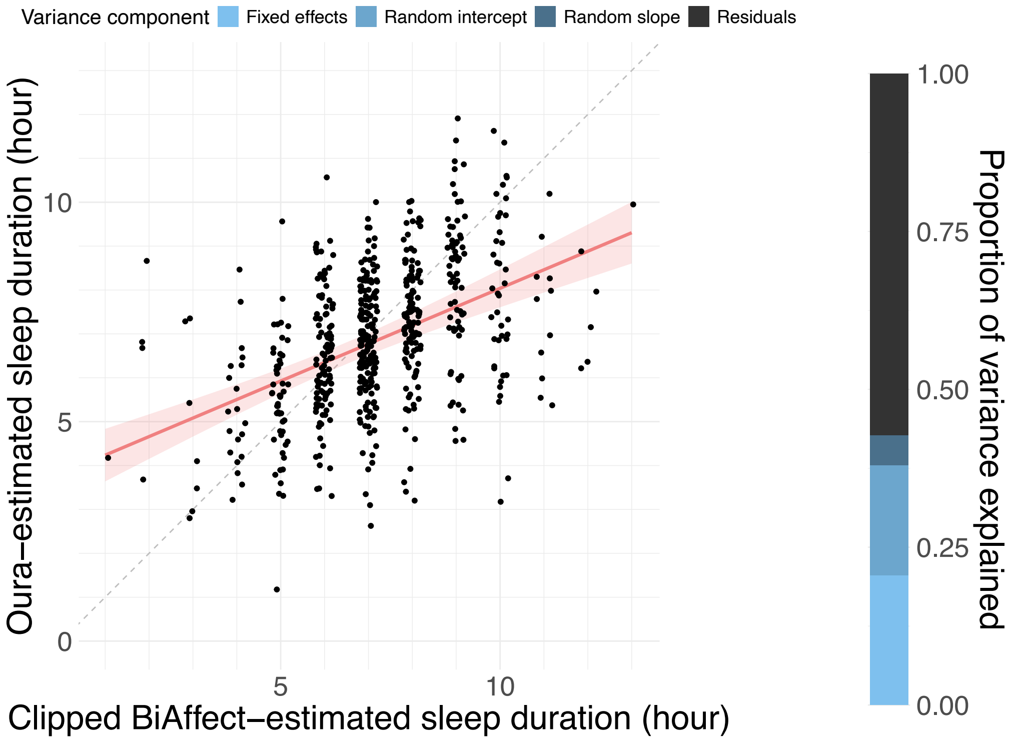

***Supplementary Figure 20: Model predictions using additional typing modalities.*** *Jittered data points are plotted over the regression line. The shaded band represents the 95% CI.*

### Supplementary Note 19 – Phase values by time zone

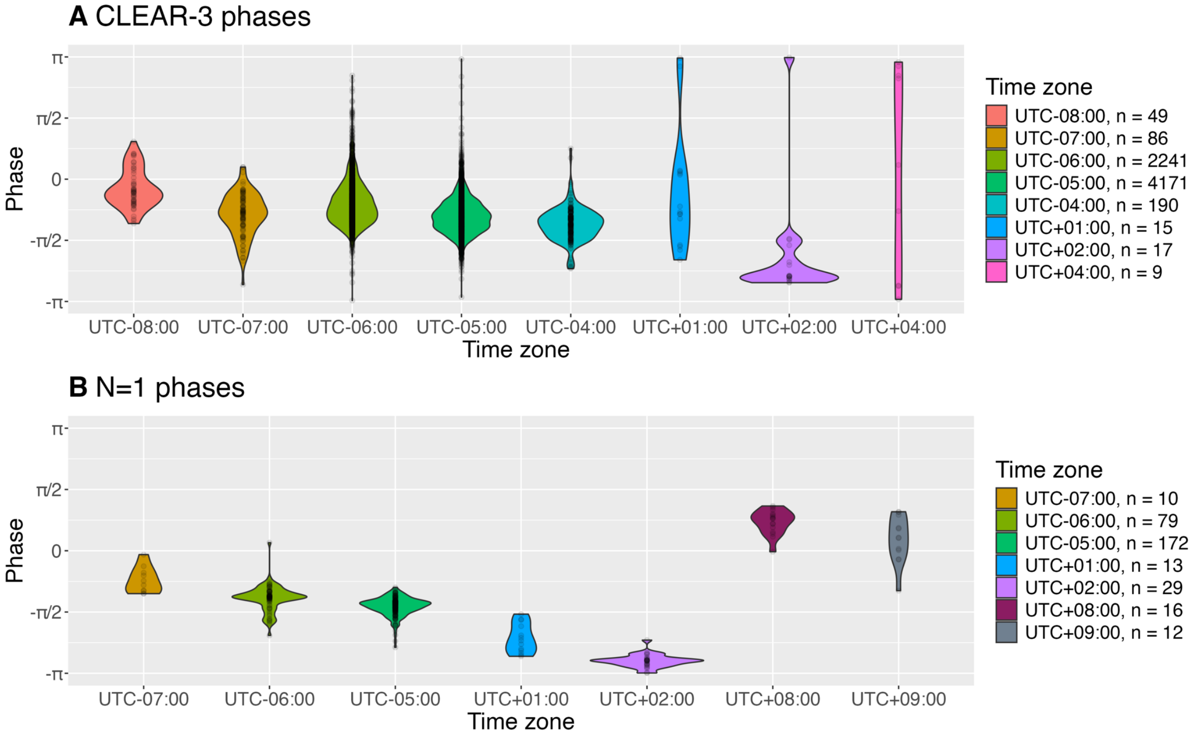

***Supplementary Figure 21: Phase by time zone. A*** *CLEAR-3 phases. Note that time zones with few observations might not provide a representative view of the population-level distribution.* ***B*** *Phases of author AL.*

Supplementary Figure 21 shows violin plots per time zone of phases derived from the BiAffect data. The method is designed to detect at which point during the day a person is most active, so the phases are expected to be different between time zones. However, day-to-day variations and chronotype will change when a person is most active, and not every day will see regular phone use, so these phases will inevitably display between- and within-person variation within a single time zone. This becomes clear from the CLEAR-3 phases, which suggest large variation within but small average differences between time zones. These average differences can be obscured when time zones lack enough data, which is especially prominent in UTC+04:00.

A slightly cleaner signal can be obtained by considering the N=1 data, which removes between-person variation due to, e.g., chronotype. Additionally, author AL, the contributor of these data, used BiAffect regularly throughout the data collection period. The result is that the differences in phase between time zones is more apparent, despite the lower number of observations per time zone.

9. Ning, E. *et al.* Smartphone-derived Virtual Keyboard Dynamics Coupled with Accelerometer Data as a Window into Understanding Brain Health: Smartphone

Keyboard and Accelerometer as Window into Brain Health. in *Proceedings of the 2023 CHI Conference on Human Factors in Computing Systems* 1–15 (ACM, Hamburg Germany, 2023). doi:10.1145/3544548.3580906.

10. Altini, M. & Kinnunen, H. The Promise of Sleep: A Multi-Sensor Approach for Accurate Sleep Stage Detection Using the Oura Ring. *Sensors* **21**, 4302 (2021).

11. Shlens, J. A Tutorial on Principal Component Analysis. Preprint at https://doi.org/10.48550/arXiv.1404.1100 (2014).

12. Cai, D., He, X., Han, J. & Huang, T. S. Graph Regularized Nonnegative Matrix Factorization for Data Representation. *IEEE Trans. Pattern Anal. Mach. Intell.* **33**, 1548–1560 (2011).

13. Vidar, E. A. & Alvindia, S. K. SVD Based Graph Regularized Matrix Factorization. in *Intelligent Data Engineering and Automated Learning – IDEAL 2013* (eds Yin, H. et al.) 234–241 (Springer, Berlin, Heidelberg, 2013). doi:10.1007/978-3-642-41278-3_29.

14. Donelli, P. New Functional Data Analysis Methods, with Applications to Spatial Transcriptomics and Neuroimaging. (2023).

15. Druijff-van de Woestijne, G. B. *et al.* Behavioural biometrics: Using smartphone keyboard activity as a proxy for rest–activity patterns. *J. Sleep Res.* **30**, e13285 (2021).

16. Otsu, N. A Threshold Selection Method from Gray-Level Histograms. *IEEE Trans. Syst. Man Cybern.* **9**, 62–66 (1979).

17. Shrivastava, D., Jung, S., Saadat, M., Sirohi, R. & Crewson, K. How to interpret the results of a sleep study. *J. Community Hosp. Intern. Med. Perspect.* **4**, 24983 (2014).
